# Resolving heterogeneity in Diffuse Large B-cell Lymphoma using a comprehensive modular expression map

**DOI:** 10.1101/2022.05.23.22275358

**Authors:** Matthew A. Care, Daniel Painter, Sharon Barrans, Chulin Sha, Peter Johnson, Andy Davies, Ming-Qing Du, Simon Crouch, Alex Smith, Eve Roman, Cathy Burton, Gina Doody, David Westhead, Ulf Klein, Daniel J. Hodson, Reuben Tooze

## Abstract

Diffuse large B-cell lymphoma (DLBCL) is characterised by pronounced genetic and biological heterogeneity. Several partially overlapping classification systems exist – developed from mutation, rearrangement or gene expression data. We apply a customised network analysis to nearly five thousand DLBCL cases to identify and quantify modules indicative of tumour biology. We demonstrate that network-level patterns of gene co-expression can enhance the separation of DLBCL cases. This allows the resolution of communities of related cases which correlate with genetic mutation and rearrangement status, supporting and extending existing concepts of disease biology and delivering insight into relationships between differentiation state, genetic subtypes, rearrangement status and response to therapeutic intervention. We demonstrate how the resulting fine-grained resolution of expression states is critical to accurately identify potential responses to treatment.

**Significance statement:** We demonstrate how exploiting data integration and network analysis of gene expression can enhance the segregation of diffuse large B-cell lymphoma, resolving pattens of disease biology and demonstrating how the resolution of heterogeneity can enhance the understanding of treatment response.

## Introduction

Heterogeneity is a characteristic feature of diffuse large B-cell lymphomas (DLBCL). Consistent subtypes have been resolved based on expression state, mutational profiles and cytogenetic features. Amongst the most significant insights has been the association of DLBCL with either germinal centre (GCB) or post-germinal centre/activated B-cell (ABC) counterparts.^1^ More recently analysis of mutation patterns in DLBCL have converged onto recurrent patterns of co-mutation that can be used to define genomic subtypes.^2–7^ A third approach widely used in clinical practice is separation by gene rearrangement status with identification of high-risk DLBCL cases based on double-hit (DH) or triple-hit (TH) rearrangements of *MYC* and *BCL2* and/or *BCL6* genes.^8–11^

Expression-based classification of DLBCL is not restricted to identification of cell-of-origin (COO) classes, with the parallel consensus cluster classification (CCC) focusing on metabolic, signalling and host response features.^12^ The latter have also been identified in separate stromal survival predictors.^13^ Furthermore, high-risk DLBCL cases have been identified based on gene expression features, learned either from patterns in Burkitt lymphoma,^14^ or based on direct similarity to cases with *MYC* and *BCL2* DH.^15^ These approaches identify overlapping sets of cases and are enriched amongst DLBCL associated with mutations of *EZH2* and *BCL2*.^2,4,5^

Single-cell expression analysis of germinal centre (GC) B-cells has provided additional insight, separating features of the two main functional populations of dark and light zones, in which B-cells undergo proliferation, somatic hypermutation and T-cell mediated selection, along with intermediate populations including those transitioning to post-GC differentiation.^16–21^ Additionally, the calculated contributions of multiple cell states, including both neoplastic B-cells and accompanying host response has been used to assign DLBCL to ecotypes;^22^ and functional expression signatures have been used to define differences in lymphoma microenvironments across both conventional COO classes and genomic DLBCL categories.^23^ However, an integrated picture unifying features of expression, mutation and rearrangement status is still developing. Here we address this issue using an approach which resolves the intrinsic structure of gene co-expression. Drawing on a broadly representative resource of close to 5000 DLBCL cases across prior studies, we demonstrate how the resultant integrated view of DLBCL expression biology enhances the informed selection of features for expression-based classification.

## Results

### An integrated gene expression context for DLBCL

To provide a comprehensive exploration of DLBCL biology from the perspective of gene expression we employed parsimonious gene correlation network analysis (PGCNA) (Supplemental Figure 1).^24^ This approach allows integration of multiple datasets across platforms and makes use of radical edge reduction to efficiently derive the configuration of large networks and optimize modularity.^24,25^ In this approach the focus for each gene is on the mostly highly correlated gene partners, hub nodes/genes in the network emerge consequent to being highly correlated partners of many other genes and the primary determinant of resolved modules is the pattern and consistency of gene co-expression in DLBCL data.

We utilised a broadly representative sample of available DLBCL datasets divided into two components. For network discovery we used gene expression derived from 14 DLBCL datasets, restricting to datasets with >50 DLBCL cases, encompassing a total of 2,505 cases.^2,12,13,26–35^ The resulting network reflects a representative sample of DLBCL expression data from multiple sources, mitigating against biases linked to case selection and platform features. Recent datasets that combine mutation and expression features, totalling 2,484 cases, were reserved for downstream analysis.^3,5,14,36,37^

The resulting DLBCL expression network resolved into 28 modules of gene co-expression, across 16,054 genes/nodes (Figure 1a, https://mcare.link/DLBCL2, Supplemental Table 1). A comprehensive gene ontology and signature enrichment was used to assess biological features associated with these modules. Representative terms were selected to identify notable expression patterns associated with each module (Figure 1b, Supplemental Figure 2, Supplemental Table 2).

**Figure 1.**
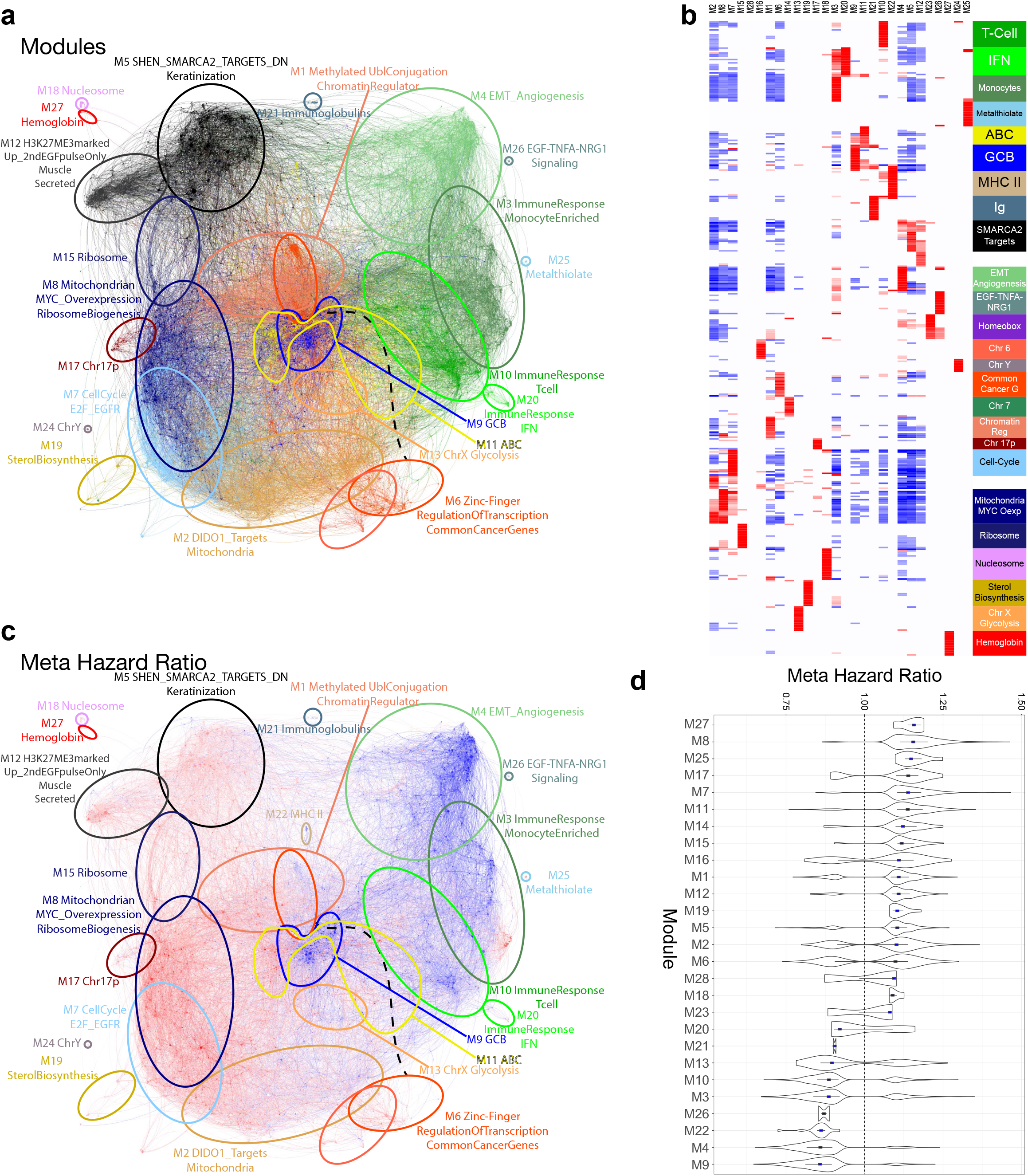
PGCNA network visualization for DLBCL. (**a**) DLBCL network with modules colour-coded, modules outlines are approximated with ellipses of same colour with module summary term indicated. Interactive versions are available at https://mcare.link/DLBCL for detailed exploration. (**b**) Separation of module signature and ontology associations is illustrated as a heatmap (filtered FDR <0.05 and ≥ 5 and ≤ 1000 genes; top 15 most significant signatures per module). Significant enrichment or depletion illustrated on red/blue scale, x-axis (modules) and y-axis (signatures). Hierarchical clustering according to gene signature enrichment. For high-resolution version and extended data see Extended Data Figure 2 and Supplemental Table 2. (**c**) Overlay of meta-hazard ratio (HR) of death across available meta-data in training datasets. Corresponding module outline approximations are illustrated as in (a). Colour scales: outcome blue (low HR - good outcome) to red (high HR - poor outcome). Interactive version available at https://mcare.link/DLBCL for detailed exploration, along with additional meta-data overlays. (**d**) Ranked module level association with HR of death. Distribution of HR associations for module genes with p-value < 0.05, along with median (blue square) and IQR.

Several features were evident. Classifier genes used for separation of ABC and GCB classes in the original COO classifier divided discretely and exclusively between two modules: M9 containing all GCB classifier genes, and M11 containing all ABC classifier genes (M11 ABC: p-value 6.95×10^-19^ and M9 GCB: 1.57×10^-11^).^38^ These two modules also capture the majority of B-cell lineage gene expression as assessed by signature enrichment (e.g. SignatureDB pan-B or Resting Blood B-cell). Thus, the fundamental separation of GCB-and ABC-related expression patterns was rediscovered as a primary feature of gene co-expression patterns in the network structure. The more numerous CCC genes divided across several modules but with significant enrichments in specific modules (M9:B-cell receptor p-value 3.27×10^-57^, M3:Host response p-value 1.64×10^-64^ and M13:Oxphos p-value 1.38×10^-05^).^12^ Additional modules separated features related to MYC function (M8), cell cycle (M7), glycolysis (M13), and sterol biosynthesis (M19). Host response/microenvironment was divided between modules related to stromal/angiogenesis (M4), T/NK-cells (M10), monocytes/macrophages (M3) and IFN responses (M20). Highly co-ordinated gene batteries were resolved for nucleosome components (M18), homeobox (M23), immediate early (M26) and metallothionein genes (M25). Several modules related to structural chromosomal regions including chr6 (M16), chr7 (M14), chr17p (M17), chrX (M13) and chrY (M24).

The network thus provides an integrated picture of gene expression in DLBCL encompassing details of lineage-specific gene expression set against a diverse backdrop of cell biology and microenvironment and resolving fine-grained patterns of gene co-expression. These are available as an extensive online resource covering gene correlation data, interactive network visualisations and downstream comparisons (https://mcare.link/DLBCL2).

### Meta-hazard ratio analysis confirms associations between biology and outcome

A meta-hazard ratio analysis merging overall survival data from training datasets can illustrate the association of module and individual gene expression with outcome across the entire network. We limited analysis to cases treated with R-CHOP immunochemotherapy. The result included the segregation of good and adverse outcome between component genes of the GCB (M9) and ABC (M11) modules (Figure 1c and d), as well as adverse outcome with Mitochondrial and MYC overexpression module (M8) and Ribosome module (M7) and good outcome linked to expression of the Stromal/Angiogenesis module (M4) which encompasses features of known survival predictors.^13^

### Resolving detailed differentiation states in GCB and ABC modules

Biological detail can be resolved at different levels of granularity in the network. An iterative analysis of gene correlation within modules can resolve the most highly related gene neighbourhoods, and the extent to which underlying information is successfully separated by such neighbourhoods can be assessed with gene signature enrichment.^24^

Assessed systematically across the network, the ABC and GCB modules were amongst those with highest neighbourhood-level information content (Supplemental Figure 3, Supplemental Table 3 and 4). Given the central importance of B-cell differentiation to lymphomagenesis,^39,40^ we focused on the relationships of the GCB and ABC neighbourhoods in more detail (Figure 2a and b, and https://mcare.link/DLBCL2). The GCB module (M9) resolved into 16 neighbourhoods (Figure 2b and c, Supplemental Table 4). These included neighbourhoods enriched for different GC B-cell subsets - LZ (M9_n2 and n6), DZ (M9_n5 and n15), CD40/NFκB responses linked to LZ B-cells (M9_n10), Intermediate-e/CCR6 memory B-cell precursor (M9_n3 and n4) and pre-memory/memory B-cells (M9_n14).^16^ A similar informative separation was observed for the ABC module which divided into 17 neighbourhoods (Figure 2b and d, Supplemental Table 4). This separated expression related to CCR6+ memory B-cell precursors (M11_n5), pre-memory B-cells (M11_n5 and n7), and plasmablast/plasma cell (M11_n6). These cell state associations overlapped with targets of key transcriptional regulators IRF4 (M11_n1, n6 and n7), BLIMP1 (M11_n7) XBP1 (M11_n6), and NFκB-response genes specific to the ABC modules (M11_n4 and n8).

**Figure 2.**
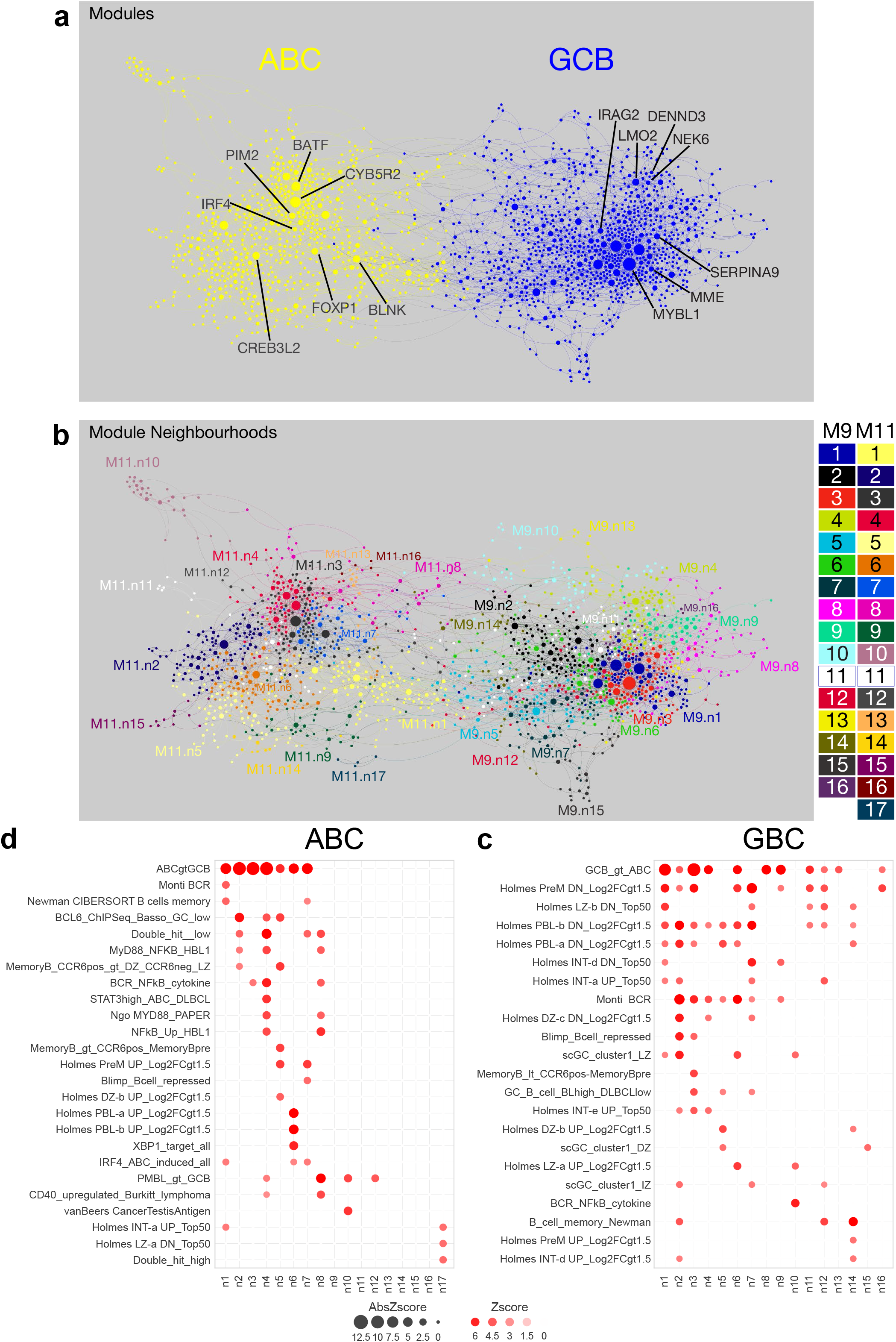
GCB and ABC module neighbourhoods provide fine-grained resolution. **(a)** Modules M9_GCB (blue) and M11_ABC (yellow) are displayed as a merged ABC/GCB specific network with node sizes reflecting degree. **(b)** The ABC/GCB specific network structure as in (a) colour coded according to gene neighbourhood membership, with colour key provided to the right. **(c)** and **(d)** Separation of neighbourhood signature and ontology associations are illustrated as a bubble plot for M9_GCB (c) and M11_ABC (d) (showing select signatures). Significant enrichment illustrated by colour and bubble size, x-axis (neighbourhoods) and y-axis (signatures) for relevant signatures. For high-resolution version and extended data see Extended Data Figure 3 and Supplemental Table 4.

The COO gene expression classifier established the paradigm for the successful application of small numbers of representative genes to separate individual DLBCL cases into distinct expression states.^38^ Genes that have a high degree of connectivity in the network may provide particularly useful information to summarise expression states of many correlated neighbours and overlapping information can be derived from different but correlated genes within network neighbourhoods/modules. The latter illustrates how different classifier gene sets can be selected to arrive at broadly similar answers for a given expression state. Many of the COO classifier genes emerged as hub nodes within the respective GCB and ABC neighbourhoods (Figure 2a). Considering the genes used in the original COO classifier, those used to classify GCB-DLBCL were all contained within the M9 module and segregated primarily between two neighbourhoods M9_n3 (*BCL6*, *MME* (*CD10*) and *SERPINA9*) and M9_n4 (*DENND3*, *ITPKB*, *LMO2* and NEK6) while *IRAG2* (*LRMP*) belonged to M9_n11. The COO ABC-classifier genes were distributed across 5 neighbourhoods in module M11 (M11_n1, n2, n3, n4 and n7), illustrating that these genes sample multiple aspects of the ABC network module (Figure 2a and Supplemental Figure 3c). We conclude that neighbourhood-level patterns of gene expression can resolve details of biology related to differentiation state and can illustrate the inter-relationship of individual genes used in targeted expression-based classifiers.

### Network gene expression patterns show consistent association with mutation state

HMRN, REMoDL-B and Reddy datasets include both gene expression and mutation data.^5,36,37^ These studies were not used in network generation. We could therefore test whether patterns of gene co-expression in the network correlated with mutations in these independent datasets. Since many mutations in DLBCL are rare we considered both the correlation of expression and mutation within individual datasets and as a consensus meta-mutation correlation across the three datasets. We summarised module-or neighbourhood-level gene expression as metagenes (module or neighbourhood expression values, MEV or NEV) and assessed the correlation of these values with the presence or absence of gene mutation, reducing mutation data for each case into a mutated/unmutated binary call for each gene with annotated putative pathogenic mutations.

Metagene expression patterns were significantly associated with mutation state (Figure 3 and Supplemental Figure 4). At module level the primary separation was into two clusters corresponding to M9_GCB and M11_ABC mutation correlations. M9_GCB correlated significantly with a wide range of mutations consistent with known associations both of COO-defined GCB-DLBCL and follicular lymphoma (FL),^3,41^ and anticorrelated with the primary mutations linked to COO-defined ABC-DLBCL. M11_ABC exhibited the reciprocal anticorrelation with mutations linked to COO-defined GCB-DLBCL and positive correlation with mutations characteristic of COO-defined ABC-DLBCL. Modules which shared mutation correlations with M9_GCB included: M13_ChrXGlycolysis, M23_Homeobox, M28_ITGB8, M6_ZincFinger, M6_Chr6; while modules with shared mutation correlations with M11_ABC included: M8_MitochondrionMYC, M18_Nucleosome, M7_CellCyle and M17_Chr17p. An additional cluster of modules focused on shared correlations with *CDKN2A* mutation including M14_Chr7 and M15_Ribosome.

**Figure 3.**
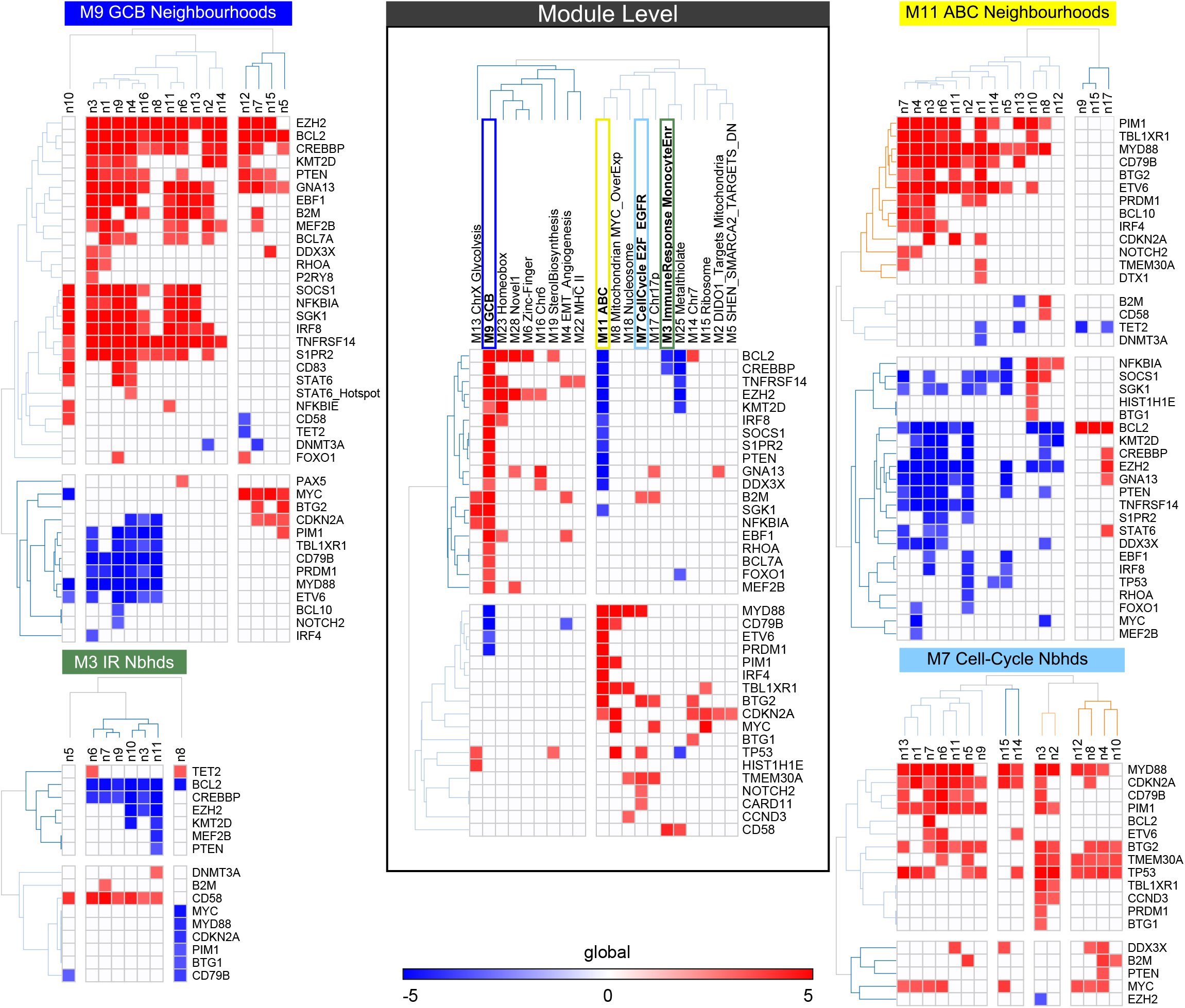
Relationship between MEV/NEVs and mutations. Association of Module/Neighbourhood Expression Values (MEV/NEV) with mutations. Shown are combined significance (Stouffer method) of p-value based on point-biserial correlations between binary mutation status and MEV/NEVs across 3 datasets (HMRN, Reddy & REMoDLB). Only mutations with p-value < 0.001, occurring in ≥ 2 datasets were retained, Z-scores for p-values > 0.001 were set to 0. Significance is shown as z-scores on a blue (significant depletion) to red (significant enrichment) scale (-5 to +5). Centre shows the relationship between MEVs and mutations. Outside shows the relationship between select NEVs and mutations.

Further subdivision was evident when modules were considered at neighbourhood-level. For instance, while most neighbourhoods in the GCB module (Figure 3 top left panel) shared correlation with a core set of gene mutations including *EZH2, BCL2* and *CREBBP* the neighbourhood-level analysis could discriminate heterogeneity for other genes. For example, neighbourhoods discretely separated between those with *SOCS1, NFKBIA, SGK1, IRF8, TNFRSF14, S1PR2, CD83*, *STAT6* mutation association and those with *MYC*, *CDKN2A* mutation association. Additionally, mutation in several genes including *BTG2, DDX3X, FOXO1, RHOA, P A* o*X*r *5STAT6* Y419 hotspot mutations associated with select patterns of GCB neighbourhood expression. Similarly, in the ABC module (Figure 3 top right panel) most neighbourhoods shared association with a common set of mutated genes including *MYD88, CD79B, PIM1, ETV6, TBL1XR1, BTG2*, *PRDM1*. However, more selective associations were evident for *BCL10, IRF4, CDKN2A, NOTCH2*, *TMEM30A*. Other M11_ABC neighbourhoods captured quite distinct mutation associations lacking positive correlation with the *MYD88, CD79B* mutation cluster and showing associations with *NFKBIA, SOCS1, SGK1*, *BTG1* or with *BCL2* mutation and features transitional to a GCB-like pattern.

Further examples of informative separation at neighbourhood-level are illustrated by the M3_ImmuneResponseMonocyteEnriched and M7_CellCycle neighbourhoods. For the M3_ImmuneResponseMonocyteEnriched module (Figure 3 bottom left panel) common association was observed for mutation in *CD58*. However, select neighbourhoods (M3_n6, M3_n8 and M3_n11) were also associated with mutation in either *TET2* or *DNMT3A*. For the M7_CellCycle module (Figure 3 bottom right panel), *TP53* mutation, was significantly associated with 12/15 of neighbourhoods which contrasted with its lack of significant association with any M8_GCB or M11_ABC neighbourhoods. Interestingly, 8/12 neighbourhoods linked to *TP53* mutation also share association with mutation of *TMEM30A*, recently identified as a tumour suppressor inactivated in DLBCL, but in contrast to *TP53* linked to a favourable response to R-CHOP therapy.^42^ Multiple M7_CellCycle neighbourhoods shared association with mutation of genes such as *MYD88, CD79B, PIM1*, *ETV6* characteristically linked to the ABC-state, while from the perspective of GCB-linked associations select M7_CellCycle neighbourhoods were associated with mutation in *MYC* and *DDX3X*.

While this analysis is limited to the sets of genes tested in targeted mutation panels, the most important known mutational features of DLBCL are included. The results identify distinct and significant associations of network derived gene co-expression patterns with driver mutations and argue that the granularity provided by the network at module and neighbourhood-level is informative of underlying tumour biology.

### Network-level gene expression patterns enhance consistent segregation of DLBCL

Given the consistent association between modules/neighbourhoods and mutation state we reasoned that network-level information may contribute to enhanced expression-based segregation of DLBCL cases. To evaluate this, we tested how recurrently discoverable the co-segregation of DLBCL cases was with different selections of features and different clustering methods. To select informative features for clustering we split all modules into neighbourhoods and considered features either at the level of individual genes or collapsing genes within neighbourhoods or modules into single values (MEV/NEVs). We then selected the most informative features at either gene, NEV or MEV level using several different approaches. The selected features were then used to cluster the DLBCL datasets (n=16, see Supplemental Table 1, DatasetInfo) using either PGCNA or consensus clustering (CC).^43^ The resultant clusterings were compared based on the extent of recurrently discoverable co-segregation of DLBCL cases. In total 33 attribute sets were examined (see Supplemental Table 5). This showed (Figure 4a): i) that selections based on PGCNA network modules with edge information (Net+Strct) outperformed ranking without edge information (Network); ii) that having only M9/M11 at neighbourhood-level (NEV_M9M11) outperformed selections using all neighbourhoods; iii) that collapsing genes per module/neighbourhood to MEV/NEVs outperformed gene-level clusterings; and iv) that using network structure and edge information (Net+Strct) to inform gene choice and collapsing genes to module/neighbourhood values (MEV/NEVs) significantly outperformed gene selection based on most variant expression (Top1000/5000). We conclude from this that use of network information can enhance the selection of attributes for consistent clustering of DLBCL cases.

**Figure 4.**
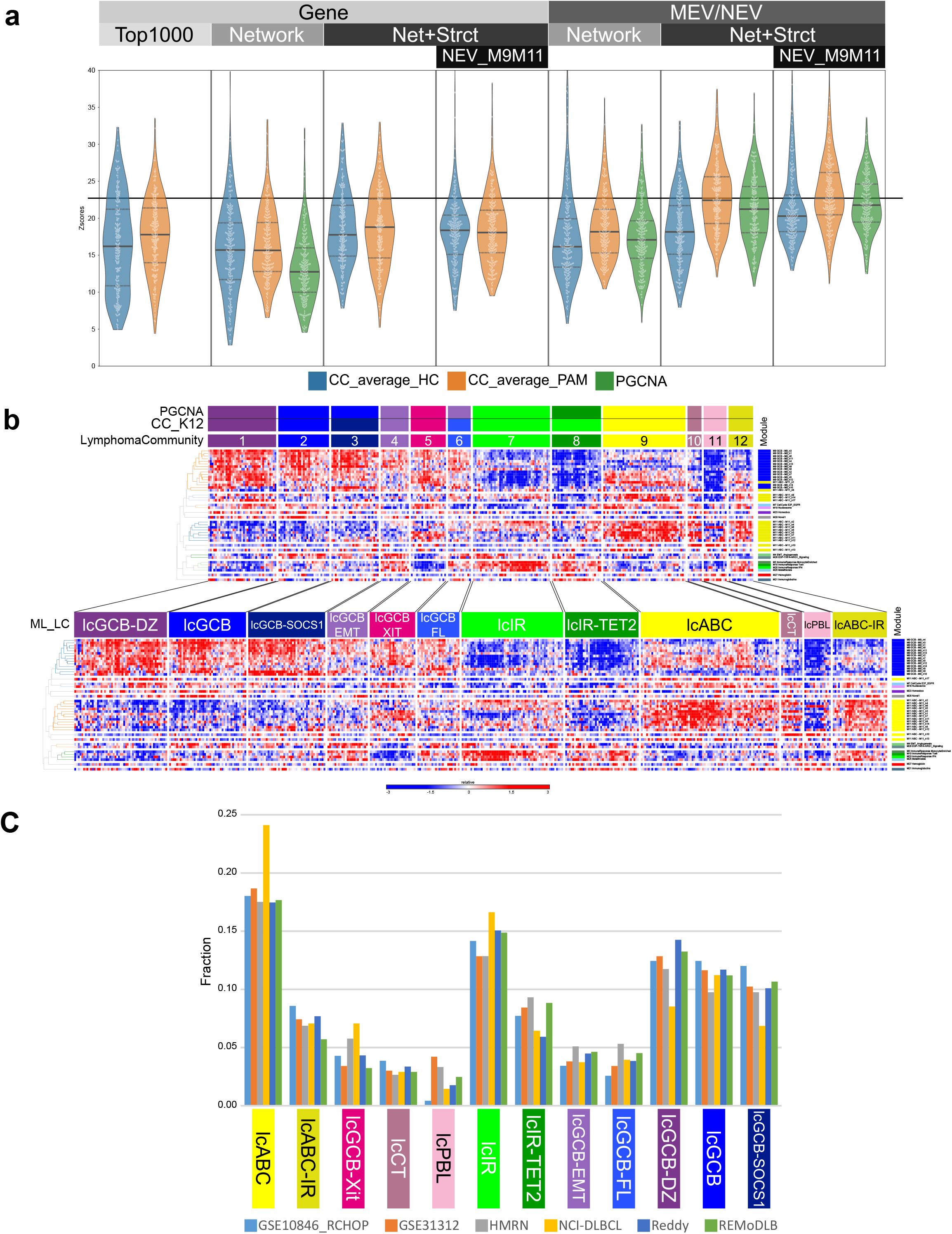
Attribute selection and building a Lymphoma Community classifier. The attributes used for clustering samples were tested using a machine learning (ML) approach (see EDF1) that assessed the recurrence of discovered clusters between clusterings of different DLBCL datasets. (**a**) displays violin plots of Z-scores showing the significance of cluster recurrence based on overlaps between clusters in different DLBCL datasets (comparing unsupervised/supervised clusterings between each pair of datasets (n=16), yielding 240 comparisons (per K level); see EDF1, ST5 and Supplemental Methods). Attributes were clustered using CC (linkage:average and method: HC/PAM) or with PGCNA. The bars at the top show the level – gene or collapsed to MEV/NEV and the attribute type – Top1000: top 1000 genes with highest median absolute deviation, Network: most variant genes per neighbourhood (SelectG_MADS), Net+Strct: most variant neighbourhoods (genes per neighbourhood selected based on network edge strength; SelectG_NEV-MADM) and Net+Strct/NEV_M9M11: most variant modules/neighbourhoods (genes per module/neighbourhood selected based on network edge strength; SelectG_NEV-MADM) with only modules M9 and M11 at the neighbourhood level. Violin plots show the median (solid line) and Q1/Q3 (dotted lines) along with each comparison (white dot, n=240) for the k=12 CC results. The horizontal black line is set at the highest median value. (**b**) using the most informative attribute set (MEV/NEV NEV_M9M11; n=41 MEV/NEV) the overlapping CC_K12 and PGCNA clusterings of the HMRN dataset formed a training dataset to build a ML classifier. The top heatmap shows the training data and the bottom shows the total HMRN dataset reclassified using the trained ML tool (ML_LC). Each row shows the expression of the displayed MEV/NEV across the samples on a blue (low) to red (high) z-score colour scale. (**c**) the fraction of each LC across the datasets classified using ML_LC.

### Network information resolves communities of lymphoma cases with shared biology

A common feature of classification based on multiple parameters is the identification of consensus cases on which various classification tools can agree when using the same features for classification, and edge cases where the classification is more ambiguous.^44^ Therefore, extending the concept of consensus or ensemble clustering,^12,45^ we focused on cases co-clustered by both CC and PGCNA methods. This identified 12 ensemble case clusters (Figure 4b), which we refer to as Lymphoma Communities (LCs). These cases were then used as a reference set to train a machine learning tool. The machine learning tool was used to reclassify all cases in the HMRN (Figure 4b) and other datasets (see Supplemental Methods for details). A similar distribution of cases falling into these communities was recovered across a range of datasets (Figure 4c and Supplemental Figure 5 & 6). These communities were named according to patterns of module expression or associated pathological/molecular features that were identified in downstream analyses.

Amongst the 12 LCs (Figure 4b, Supplemental Figure 6) two communities identified cases dominated by ABC module gene expression which were distinguished by differences in immune response-related features as immune response poor (_lc_**ABC**) or immune rich (_lc_**ABC-IR**). Cases with plasmablastic features (_lc_**PBL**) were distinguished by expression of XBP1 targets and cancer testis antigens. A distinct subset of cases with ABC-related features shared the expression of cancer testis antigens (_lc_**CT**). Amongst cases with weak B-cell patterns, two communities were dominated by immune response modules (_lc_**IR** and _lc_**IR-TET2 -** distinguished by *TET2* mutation enrichment, see below). Six communities were characterised by expression of multiple GCB-related neighbourhoods. These included a community with GCB features and distinct stromal/EMT-related gene expression (_lc_**GCB-EMT**) and a community with mixed GCB and ABC expression features along with cell cycle, MYC, and sterol biosynthesis modules, suggestive of transition from GCB to an ABC/post-GC state (_lc_**GCB-Xit**, Supplemental Figure 6). Similar proliferation and growth-related module expression were combined with polarised high GC DZ and low GC LZ neighbourhood expression, along with low CD40/NFκB and MHC-II genes in a GCB DZ-like community (_lc_**GCB-DZ**). The remaining three GCB communities were distinguished by different patterns of GCB neighbourhood expression, wider network features and other associations in downstream analyses (_lc_**GCB** - most canonical GCB-like, _lc_**GCB-FL** - underlying FL diagnosis, _lc_**GCB-SOCS1** – selective SOCS1 mutation association).

The combination of overview and gene-level granularity provided by the network afforded a further means to assess relationships between the resolved communities across all network genes or those specific to the GCB/ABC differentiation state (Supplemental Figure 7 and 8). This is exemplified at the level of GCB and ABC neighbourhood genes, where the _lc_GCB-Xit community straddles both key GCB and ABC features and contrasts with the more discrete patterns of other communities such as _lc_GCB-DZ and _lc_ABC (Supplemental Figure 7). At the whole network level, wider differences in gene expression within modules are further illustrated as seen for the comparison between _lc_GCB-FL and _lc_GCB (Supplemental Figure 8). Thus the 12 DLBCL LCs reinforced the significance of segregation into ABC and GCB expression patterns while distinguishing heterogeneity within these broad categories.

### DLBCL communities link to mutation patterns

An important test of the DLBCL LCs was whether these also showed significant association with mutation state. To address this we analysed the enrichment of mutations in DLBCL communities by integrating enrichment p-values derived across the individual HMRN, REMoDL-B and Reddy datasets.^5,36,37^ At a p-value threshold <0.01 (in ≥ 2 datasets) the communities showed significant and distinct associations with mutation patterns. These separated in concordance with the expression states between mutational features linked to ABC, GCB, and host/immune response characteristics (Figure 5a). Thus, _lc_ABC and _lc_ABC-IR shared association with *MYD88, CD79B, PIM1, PRDM1*, *CDKN2A* and differed in association of with *ETV6, TBL1XR1* and *IRF4* mutations. The ABC-related _lc_CT community was selectively enriched for *MYD88* and *PIM1* mutations and was additionally enriched for *BCL10, TP53*, *MEF2B* mutations. These patterns separated most distinctly from _lc_GCB and _lc_GCB-DZ. These shared enrichment for *BCL2*, *EZH2*, *DDX3X*, *IRF8*, *PTEN, TNFRSF14*, *GNA13* mutation. GCB-DZ was additionally enriched for *CREBBP*, *KMT2D*, *BTK, POU2F2, S1PR2, FOXO1*, *MYC, MEF2B* mutation, while _lc_GCB was enriched for *CARD11, B2M, EBF1, MSH6, RHOA*, *TET2*, *SGK1* mutation. _lc_GCB-FL shared association with *CREBBP, KMT2D, TNFRSF14* but was distinguished by association with *STAT6* mutations including significant enrichment of the *STAT6* Y419 hotspot mutation. _lc_GCB-EMT shared enrichment of *SGK 1* and *TNFRSF14* with _lc_GCB but otherwise lacked distinct associations. _lc_GCB-SOCS1 and _lc_GCB-Xit differed from other GCB-related expression groups. _lc_GCB-SOCS1 associated with *SOCS1, NFKBIA, SGK1, BTG1, GNA13, CD83, NFKBIE*, *ZFP36L1* and *STAT6* but not the *STAT6* Y419 mutation hotspot, while _lc_GCB-Xit showed a further distinctive pattern with selective enrichment for *CD70, CCND3*, *BTG2* mutations. While _lc_IR only displayed anti-correlations with mutation state likely reflecting the diluting effect of host response components, _lc_IR-TET2 cases were selectively associated with *TET2* mutations.

**Figure 5.**
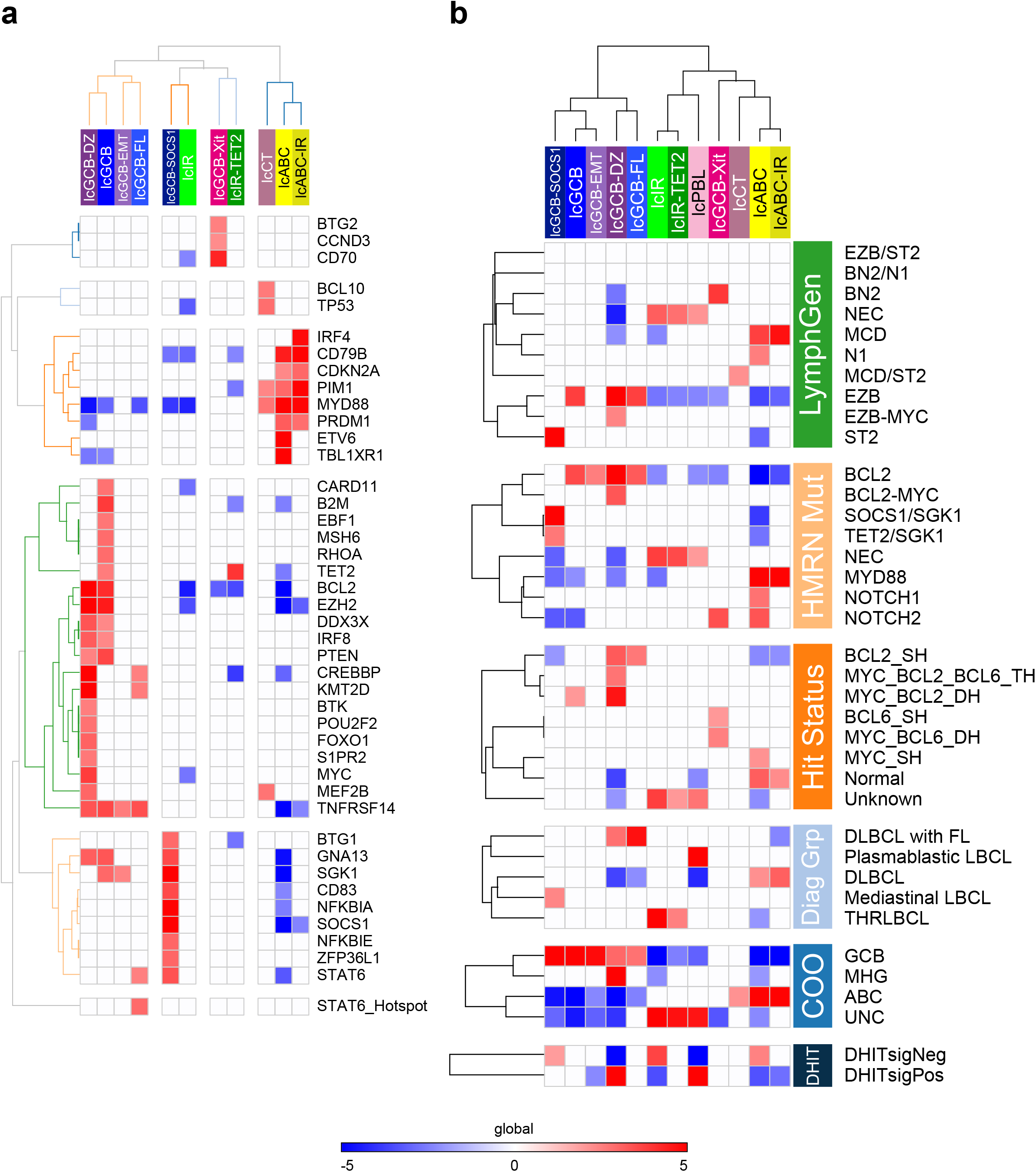
Lymphoma Communities (LC) have distinct mutational and rearrangement associations. Association of LC with mutation and rearrangement status. (**a**) The differential enrichment of gene mutations across LC integrated across three datasets. Shown is combined significance (Stouffer method) of LC enrichment/depletion of mutations as z-scores on a blue (significant depletion) to red (significant enrichment) scale (-5 to +5). X-axis shows hierarchically clustered LC and y-axis gene symbols. Only mutations with p-value < 0.01, occurring in ≥ 2 datasets were retained, Z-scores for p-values > 0.01 were set to 0. (**b**) Significance of enrichment/depletion of LC with LymphGen, HMRN mutational group (PMID: 32187361), Hit-status (rearrangement status for MYC, BCL2 and BCL6 indicating single hit (SH), double hit MYC_B-CL2-DH or MYC_BCL6-DH, or triple hit MYC_BCL2_BCL6-TH), Diagnostic-Group, cell-of-origin/MHG and DHITsig assignments in the HMRN dataset. X-axis hierarchically clustered LC, against y-axis clustered within each group. Z-scores with p-value > 0.05 were set to 0.

The observed associations between LCs and mutations resembled the patterns in recent mutation-based classifications of DLBCL. We therefore assessed the relationship between LCs and mutational classes assigned in HMRN data with either the LymphGen,^46^ or the HMRN classification (Figure 5b).^5^ These classifications though independently derived,^4,5^ are largely concordant,^46^ and recapitulate the features of the C1-C5 Harvard classification.^2^ The LymphGen MCD class was significantly associated with _lc_ABC and _lc_ABC-IR, while _lc_CT was associated with cases with ambivalent MCD/ST2 calls. LymphGen N1 showed enrichment in _lc_ABC while BN2 was significantly and selectively associated with _lc_GCB-Xit. EZB was most significantly enriched amongst _lc_GCB, _lc_GCB-DZ and GCB-FL, while EZB-MYC+ was selectively enriched amongst _lc_GCB-DZ. ST2 cases mapped selective on to _lc_GCB-SOCS1. Similar patterns of association were evident when considering the associations of LCs with the independent but related HMRN mutational classification. We conclude that the mutation-based subdivision of DLBCL in LymphGen and related classifications overlaps significantly with LCs derived independently from gene co-expression patterns.

### Lymphoma communities underline distinctions between double-hit lymphomas

In DLBCL the occurrence of double-or triple-hit with rearrangement of *MYC* and *BCL2* (MYC_BCL2_DH) or *MYC* and *BCL6* (MYC_BCL6_DH), or *MYC*, *BCL2* and *BCL6* (MYC_BCL2_BCL6_TH) identifies high-risk disease.^8–11^ The relationships of these rearrangement-based categories to gene expression and mutation features is relatively clear-cut for MYC_BCL2_DH, but remains less well-defined for MYC_BCL6_DH.^47–49^ We therefore tested the association of rearrangement status and LCs in the HMRN cohort. _lc_GCB-DZ was highly enriched for MYC_BCL2_DH and MYC_BCL2_BCL6_TH cases (Figure 5b). In contrast, a more modest enrichment of MYC_BCL2_DH cases was evident in _lc_GCB, while _lc_GCB-FL was selectively enriched for BCL2_SH. _lc_GCB-Xit was selectively enriched for MYC_BCL6_DH and BCL6_SH status but was not associated with any combination of *BCL2* rearrangement, while _lc_ABC was enriched for cases with MYC_SH.

We further assessed whether the LCs were significantly associated with morphological diagnosis, COO class and molecular high grade (MHG) or double-hit signature (DHITsig) status.^14,15^ Two communities, GCB-FL and to a lesser extent GCB-DZ were linked to concurrent or previous diagnosis of underlying FL supporting a primary separation of disease biology in transformed FL cases (Figure 5b). _lc_PBL was selectively enriched for plasmablastic lymphoma morphological diagnosis, _lc_IR and _lc_IR-TET2 for T-cell histiocyte rich large B-cell lymphoma and _lc_GCB-SOCS1 for primary mediastinal large B-cell lymphoma. At gene expression level the LC communities recovered appropriate enrichments of GCB, ABC and unclassified cases. _lc_GCB-DZ was selectively enriched for cases designated as MHG and cases that were DHITsig positive. Notably DHITsig differed from MHG in also showing significant enrichment amongst _lc_PBL. We conclude that the DLBCL LCs separate meaningfully in relation to underlying rearrangement status, morphological diagnosis, previous expression-based classifiers of high-risk disease and the presence of underlying FL.

### Lymphoma communities have prognostic significance in R-CHOP treated cases

Given the fact that LCs segregate cases with common pathological features, we assessed whether LCs showed significant and reproducible survival differences. We addressed this across 6 datasets encompassing sufficient cases treated with R-CHOP chemo-immunotherapy.^3,5,13,29,36,37^ Across multiple datasets the DLBCL community structure separated risk (Figure 6) albeit with variation evident between cohorts most notably for _lc_GCB-DZ and _lc_CT. Most adverse risk, in terms of meta-HR across the 6 datasets, was observed for assignment of cases to _lc_PBL, followed by _lc_ABC-IR, _lc_GCB-Xit, _lc_ABC and _lc_IR-TET2. Intermediate and variable risk was observed for _lc_CT and _lc_GCB-DZ. At the other end of the spectrum particularly good risk was associated with _lc_GCB-FL, _lc_GCB-SOCS1, _lc_GCB and _lc_GCB-EMT. We conclude that the refined break down of DLBCLs in the 12-fold LC structure has potential utility to refine the separation of risk groups, while remaining aligned to the concept of the COO differentiation state.

**Figure 6.**
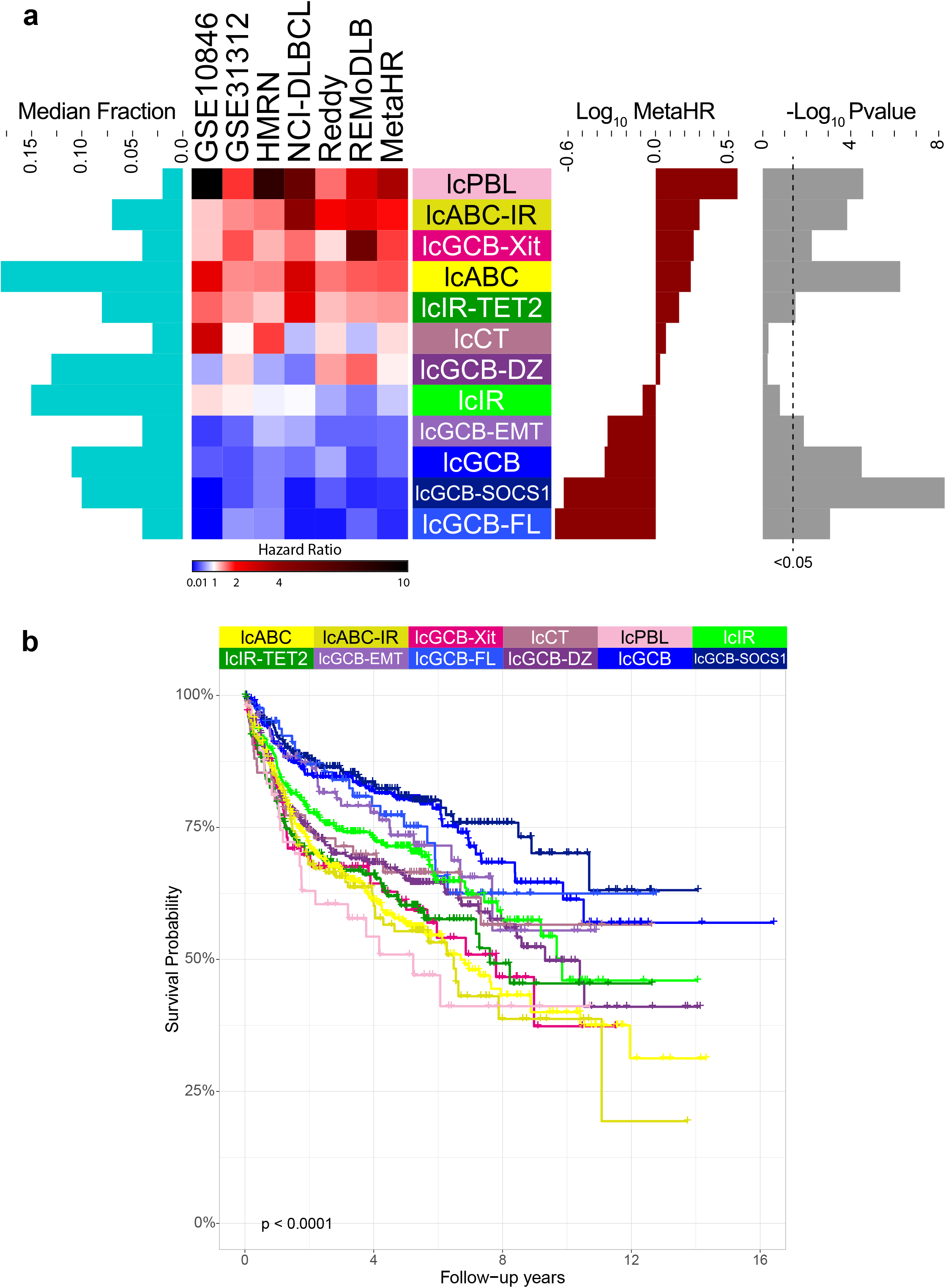
Lymphoma communities are significantly associated with overall survival. (**a**) Lymphoma Communities (LC) across the indicated datasets show consistent associations for hazard ratio (HR) of overall survival for R-CHOP treated patients. Heatmaps shows HR on a blue (good) to red (poor) colour scale across 6 datasets ordered by metaHR (from Cox proportional hazards regression; ML_LC LC p-value as the explanatory variable). The left chart shows the median fraction size of each LC across the 6 datasets. The right charts show the Log10 MetaHR (red) and -Log10 p-value (grey) with p<0.05 indicated by a dashed-line. (**b**) Kaplan-Meier plots of overall survival for R-CHOP treated patients across the 6 DLBCL datasets for shown LC; p-value from log-rank test.

### Lymphoma communities show distinct responses to RB-CHOP

The REMoDL-B trial recently reported on extended follow-up identifying a benefit for proteasome inhibitor bortezomib with R-CHOP in cases classified as ABC-DLBCL.^50^ We were therefore interested to test whether LCs could provide further insight into responses to RB-CHOP. For overall (OS) and progression-free (PFS) survival the LCs were significantly separated overall in the R-CHOP arm of the trial. In the RB-CHOP arm the significance of separation for the LCs declined for both OS (R-CHOP p=0.0013, RB-CHOP p=0.11) and PFS (R-CHOP p=0.002, RB-CHOP p=0.02) (Figure 7). While there are inherent limitations in *post hoc* analysis and the impact of 12-fold classification on case numbers, we were interested to further compare the response for R-CHOP and RB-CHOP arms of the trial for each LC (Supplemental Figure 9 and 10). While some indication of improved responses for the RB-CHOP arm was evident, for example for _lc_PBL case that achieved a 90%+ OS in the RB-CHOP treated arm (OS p=0.04, PFS p=0.014), surprisingly, no difference in outcome was observed for _lc_ABC or _lc_ABC-IR between R-CHOP and RB-CHOP arms.

**Figure 7.**
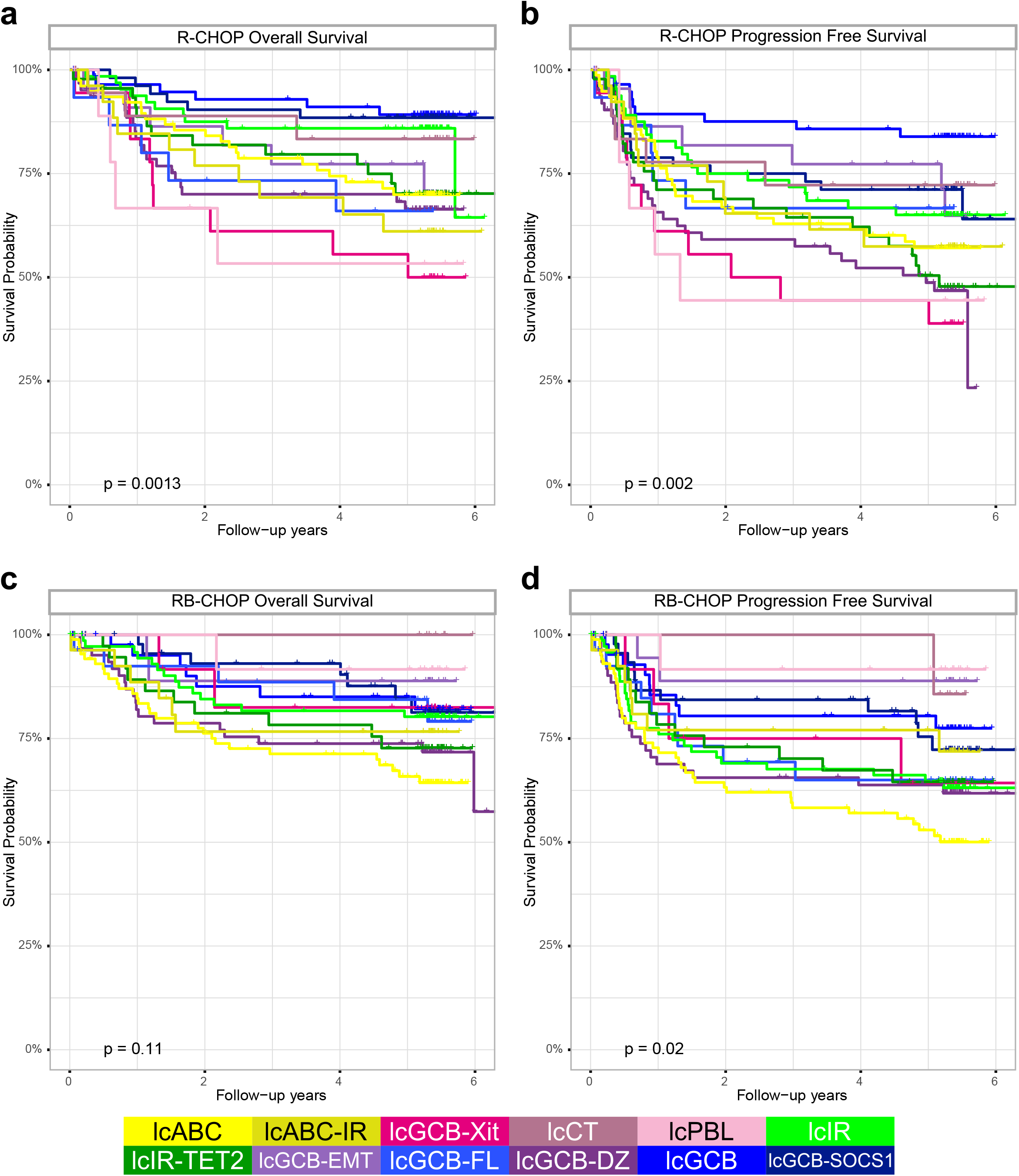
lcPBL and lcCT show improved survival with addition of bortezomib. Shows Kaplan-Meier plots of survival for the indicated Lymphoma Communities (LC) within the REMoDLB dataset split by treatment (R-CHOP or with addition of bortezomib; RB-CHOP) and survival (overall-survival:OS, progression-free-survival:PFS). (**a**) R-CHOP OS, (**b**) R-CHOP PFS, (**c**) RB-CHOP OS and (**d**) RB-CHOP PFS. P-value from log-rank test.

### Bortezomib response in ABC cases with variant expression features

The absence of survival difference between the two arms of the trial for cases classified as _lc_ABC contrasted with the apparent benefit of RB-CHOP amongst cases classified as ABC-DLBCL using the trial classifier (_T_ABC). We therefore examined the relative distribution of _T_ABC cases across the LC assignments in REMoDL-B (Figure 8A). Of the 249 _T_ABC cases 120 were also assigned to _lc_ABC and 47 were assigned to _lc_ABC-IR (together 67% of _T_ABC). These intersects included cases with most characteristic ABC expression patterns and showed no significant survival separation between RB-CHOP and R-CHOP arms of the trial. In contrast there was a suggestion of benefit for RB-CHOP over R-CHOP in _T_ABC cases assigned to other LCs. Thus, we conclude that response to RB-CHOP rather than being associated with the most typical of ABC expression patterns is instead associated with cases that combine some ABC expression features with additional features that lead to other (non-ABC) assignments in the LC structure.

**Figure 8.**
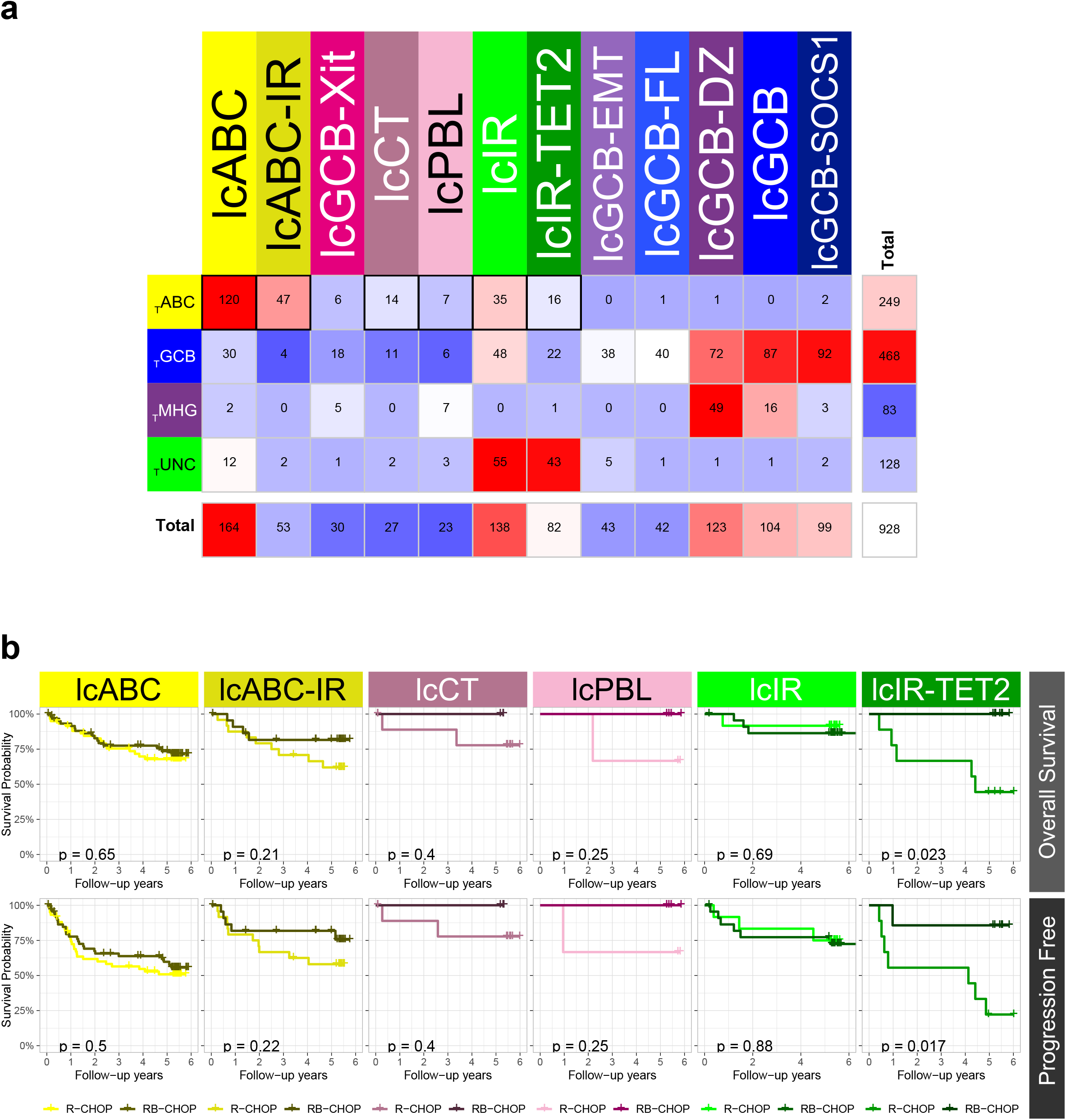
High confidence ABC does not show significant survival advantage with addition of bortezomib. (**a**) shows the overlap between REMoDLB trial COO assignments (rows) and Lymphoma Communities (columns). The samples used in part b have a black border. (**b**) Shows Kaplan-Meier plots of survival for Lymphoma Community overlap subsets of the TABC group. Each plot compares the treatments: R-CHOP (lighter-line) and RB-CHOP (darker-line) with overall-survival (top row) and progression-free-survival (bottom row). P-value from log-rank test.

## Discussion

Heterogeneity is a dominant feature of DLBCL biology. This has led to partially intersecting taxonomies. The most widely accepted of these are classification based on *MYC* and *BCL2* rearrangement status, COO classification based on gene expression patterns, and the recent mutational classifications defined in the LymphGen, Harvard and HMRN classifications.^1–4^ However, there remains uncertainty about the inter-relationships between rearrangement, expression and mutation-based classifications.

Here we have used a framework of consistent patterns of gene expression and an inclusive, correlation-centred approach to address these questions from a different perspective. Our analysis learned from 2,500 cases drawing on broadly representative contributions of the DLBCL research community and was tested in nearly 2,500 additional cases from recent studies.^2,12,13,26–35^ The network generated using PGCNA derives modules of co-expressed genes that reflect inherent features of DLBCL expression data and reinforces the central importance of the subdivision between ABC and GCB states. The analysis illustrates how individual genes relate to each other within this context. Exemplifying this, the original COO classifier genes segregate discretely between the two primary modules linked to features of B-cell lineage, and many of the classifier genes and those in alternate COO classifiers, emerge as hub-nodes with high information content in the network.^34,38^ The pattern of gene correlation at network level, illustrates how for any module or neighbourhood closely correlated genes could be selected to report similarly on the expression state.

When applying the network to expression-based classification, we have found that using correlation patterns captured in the network structure to select genes which are then collapsed into metagenes reflecting module-or neighbourhood-level expression features, as opposed to selecting highly variant genes and using these at gene-level, adds significant value to the reproducibility of classification. For DLBCL, while the prevailing models for COO classification enhanced with assessments such as MHG/DHITsig status or host response features already provide significant value to identify subsets of disease,^14,15,22,23,34,38,44^ our analysis illustrates that the integration of network features can discriminate expression patterns that are not captured in previous approaches. In terms of clinical practice, the LC structure could potentially be developed for case-by-case application, but that has not been the intent of the current study.

Our approach is distinct from and complimentary to other recent studies of DLBCL expression biology such as the Ecotyper that highlight the importance of distinct patterns of host response in DLBCL biology.^22,23^ These concepts build from earlier work identifying features of host response that are linked to good outcome.^12,13^ The importance of immune and stromal response features is underlined by the contribution that the related network modules make to distinguishing subsets of cases. An interesting example is the link between specific patterns of the M3_ImmuneResponseMonocyteEnriched neighbourhoods and mutations in *TET2* and *DNMT3A*. Indeed, an association between COO unclassified DLBCL and *TET2* mutation enrichment has previously been identified,^3^ and our analysis extends this to refine the subgroup of immune response-rich DLBCL with *TET2* mutation association. *TET2* inactivation can contribute to lymphomagenesis in a B-lineage intrinsic fashion in murine models. However, *TET2* inactivation contributes as an early event in haematopoiesis in these models and mutations in *TET2* shared between DLBCL and subsequent myelodysplasia/leukaemia in individual patients have been reported.^51–53^ Similar to *TET2*, *DNMT3A* mutations are also common features of clonal haematopoiesis/clonal cytopenia of undetermined significance.^54–56^ In angioimmunoblastic T-cell lymphoma *TET2* and *DNMT3A* mutation have been identified as early events shared with both concurrent clonal haematopoiesis and subsequent DLBCL or other haematological malignancy.^57^ It will therefore be interesting to establish to what extent the expression patterns of _lc_IR-TET2 identifies patients with DLBCL who have associated clonal haematopoiesis and whether the mutations in such DLBCL derive exclusively from neoplastic B-cells, or potentially in some cases from components of the host response.

We demonstrate that the network-based approach can facilitate the recognition of groups of cases based on expression state that overlap with mutational classifications, rearrangement status and the presence of underlying FL. In the context of cases with underlying FL the cases separate between those with balanced GCB neighbourhood-level expression patterns and excellent prognosis on R-CHOP treatment and those with a dominant DZ-like expression profile with a poorer prognosis. The _lc_GCB-DZ cases are also enriched for high-risk features such as *MYC* and *BCL2* rearrangement and MHG and DHITsig expression profiles.^8,10,14,49,58^ In contrast the good risk GCB-FL category is significantly enriched for *STAT6* mutations including the Y419F hotspot which has been characterised as a distinctive feature of a subset of FL.^41,59–62^ The GCB-FL category is in many ways consistent with recent studies showing that DLBCL with concurrent or underlying FL at diagnosis is not necessarily associated with adverse prognosis.^63,64^ Together the GCB-DZ and _lc_GCB-FL categories argue for divergent patterns of FL evolution that are distinguished on the one hand by *MYC* rearrangement and related expression features for _lc_GCB-DZ and on the other for _lc_GCB-FL a link with underlying *STAT6* mutation.

The treatment landscape of DLBCL is rapidly evolving and introduction of polatuzumab vedoitin targeting CD79b is changing frontline therapy.^65^ However, it remains of interest to explore refined separation of expression state in R-CHOP based trial data. Recently the longer-term follow-up in the REMoDL-B trial has been reported identifying a significant survival benefit in the trial-classified ABC or MHG cases.^50^ Our analysis demonstrates that the apparent response to RB-CHOP in the ABC-DLBCL subset observed in the trial classifications is not associated with the set of cases that are identified both by the trial and the LC classification as ABC. This overlap includes cases with the most typical ABC expression patterns. Instead, the response appears to reside in cases with variant features related to host response or differentiation state. We acknowledge that the *post hoc* subdivision of trial classified cases into multiple subgroups raises substantial caveats. However, a notable feature is that cases identified as plasmablastic (_lc_PBL) and treated with RB-CHOP in the REMoDL-B trial show a favourable outcome. This is notable because _lc_PBL cases in other case series have the worst outcome overall when treated with R-CHOP, which has until recently been widely used for such cases.^66^ Our analysis of the REMoDL-B data supports the arguments put forward from other studies that bortezomib-containing regimens should be considered in DLBCL with plasmablastic features.^66–70^

We conclude that our network-based approach yields an encompassing map of DLBCL tumour biology and illustrates how data integration and network analysis can refine expression-based stratification of DLBCL. Our analysis supports the argument that such refined stratification is needed to accurately identify treatment responses even within existing molecular subtypes.

## Methods

(See also Supplemental Methods)

### Expression datasets

Thirteen expression datasets were used for PGCNA (GSE4475, GSE4732, GSE10846, GSE12195, GSE19246, GSE22470, GSE31312, GSE32918, GSE34171, GSE53786, GSE87371, GSE98588, Monti).^2,12,13,26–35^ For validation 4 datasets were used: HMRN (GSE181063), NCI (NCICCR-DLBCL), REMoDLB (GSE117556) and Reddy (EGAS00001002606), (Supplemental Table 1).^3,5,13,29,36,37^ For RNA-seq datasets (NCI, Reddy) count data was processed using DESeq2 v1.22.2 with VST-normalised data used for analysis. Probes were re-annotated (http://mygene.info) ambiguous mappings were manually assigned. Datasets were quantile normalised (Python qnorm) and probe sets merged (median value for probe sets with Pearson correlation ≥0.2 and maximum value for those with correlation <0.2).

### Network generation

Discovery datasets (GSE10846 split into CHOP/RCHOP-treated) were processed using PGCNA2 (https://github.com/medmaca/PGCNA/tree/master/PGCNA2), retaining the top 70% most variant genes present in ≥50% of the datasets, carrying out 1000 Leidenalg clusterings and selecting the best using Scaled cluster enrichment scores. The resulting network contained 16,054 genes (44,730 edges) split into 28 modules (Supplemental Table 1). The network was visualised using Gephi (version 0.9.2), and interactive HTML5 web visualisations exported using the sigma.js library. Interactive networks are at https://mcare.link/DLBCL2.

### Network analysis

#### Clustering samples

See supplemental methods: Before clustering, we split each module into submodules/neighbourhoods and collapsed the genes within these to single values (MEV/NEVs). The most informative genes/MEV/NEVs were selected using several approaches, and used to cluster the DLBCL datasets using two different approaches PGCNA/ConsensusClustering (CC)^1^. We used cluster results to explore how recurrently discoverable the DLBCL communities were, allowing the selection of the most informative attributes for clustering. The best results for the HMRN dataset were combined between PGCNA/CC to generate ensemble Lymphoma Communities (LC). These were used to train a machine learning tool to recover the LCs in every dataset.

#### Neighbourhoods

Each of the 28 modules was sub-clustered to create neighbourhoods. The existing PGCNA edge file was split at the module level and then clustered 5,000 times using Leidenalg. The best clustering (based on modularity score) for each module was retained. Multiple different runs converged onto the same answer. The neighbourhoods are detailed in Extended Data Figure 3.

#### Module Expression Values and Lymphoma communities

Genes per module/neighbourhood were collapsed down to single values: within each dataset, which vary in available genes, the genes per module/neighbourhood were ranked by gene_strength (sum of genes edges/correlations within its module). Representative genes were selected and converted into a MEV or NEV by:

1. Per module/neighbourhood select top 10 genes based on ranks.
2. Per gene, standardize (z-score) the quantile-normalized log_2_ expression data.
3. Per sample (patient) calculate the median of the 10 z-scores to give a MEV/NEV.

### Lymphoma Community machine learning tool

Clustering results for HMRN were merged by selecting significant overlaps (p-value < 0.0001) that form a community in either CC/PGCNA containing > 5 samples (Supplemental Figure 1 & Supplemental Methods). This generated a high confidence set of LCs (n=298) that formed a training dataset (Figure 4b top) that was used to train a machine learning (ML) tool to recover the LC in other datasets. The training dataset was split using the python Scikit-learn test_train_split function, stratifying on the LC class label, to give class-balanced randomised training (n=238) and validation (n=60) datasets. The validation dataset was set aside to test the final selected model. The training data was used to carry out stratified 5-fold cross-validation across 7 different machine learning methods. In total 4,802 parameters were tested across the ML tools, scoring with the Matthews correlation coefficient (MCC). The best 3 models, using their optimal parameters, were combined using soft voting to give a model with mean MCC of 0.89 across the 5-folds. This model was then tested on the unseen validation data (n=60) with MCC of 0.87. This final model, termed ML_LC, was retrained using all 298 training samples and was used for all subsequent classifications, including the HMRN dataset (Figure 4b bottom).

### Survival analysis

Right-censored survival data, where available, was analysed using Survival library for R. The expression of each gene (as z-score) or the ML_LC *p*-value was used as a continuous variable in a Cox Proportional Hazards model and the ML_LC community for a Kaplan-Meier estimator (using merged survival data). Meta-analysis across datasets was conducted by fitting a fixed-effect model to hazard ratios, weighted by dataset size.

### Mutation analysis

Analysis was carried out for HMRN (188 genes/431 samples), REMoDLB (70 genes/400 samples) and Reddy (150 genes/624 samples) datasets. Mutations were converted to a binary matrix for downstream analysis. For each dataset the point-biserial correlations were calculated between all pairs of mutated gene and MEV/NEV. The resulting correlation *p*-values were converted to z-scores (python stats.norm.ppf) to convey the ±correlation along with its significance. Significance of overlap between mutations and LC was calculated using hypergeometric testing within each dataset. To generate meta results MEV/Community mutation analysis p-values were combined using the Stouffer’s Z method.

### Data Availability Statement

The underlying primary datasets are available at the indicated data source (see above under Expression Datasets). All resulting gene correlation data, module and neighbourhood gene lists, and signature/ontology enrichments are available at https://mcare.link/DLBCL2.

### Code Availability Statement

The DLBCL LC classifier and networks are available at https://mcare.link/DLBCL2. PGCNA is available at https://github.com/medmaca/PGCNA/tree/master/PGCNA2 All other code is available on request.

## Data Availability

All data produced are available at https://mcare.link/DLBCL2

## Acknowledgements

This work was supported by Cancer Research UK program grant (C7845/A17723 and C7845/A29212) (M.C, G.D., D.W, and R.T). HMRN is supported by Cancer Research UK program grant A29685 (D.P., S.C., A.S., E.R.). D.J.H. was supported by a fellowship from Cancer Research UK (CRUK) (RCCFEL∖100072) and received core funding from Wellcome (203151/Z/16/Z) to the Wellcome-MRC Cambridge Stem Cell Institute and from the CRUK Cambridge Centre (A25117). D.J.H is supported by the National Institute for Health and Care Research (NIHR) Cambridge Biomedical Research Centre (BRC-1215-20014). R.T., G.D., E.R., A.S., D.P., D.W. are supported by the National Institute for Health and Care Research Leeds Biomedical Research Centre. The views expressed are those of the authors and not necessarily those of the NIHR or the Department of Health and Social Care. For the purpose of Open Access, the authors have applied a CC BY public copyright licence to any Author Accepted Manuscript version arising from this submission.

