## Supplemental Figure 1 for "Resolving heterogeneity in Diffuse Large B-cell Lymphoma using a comprehensive modular expression map"

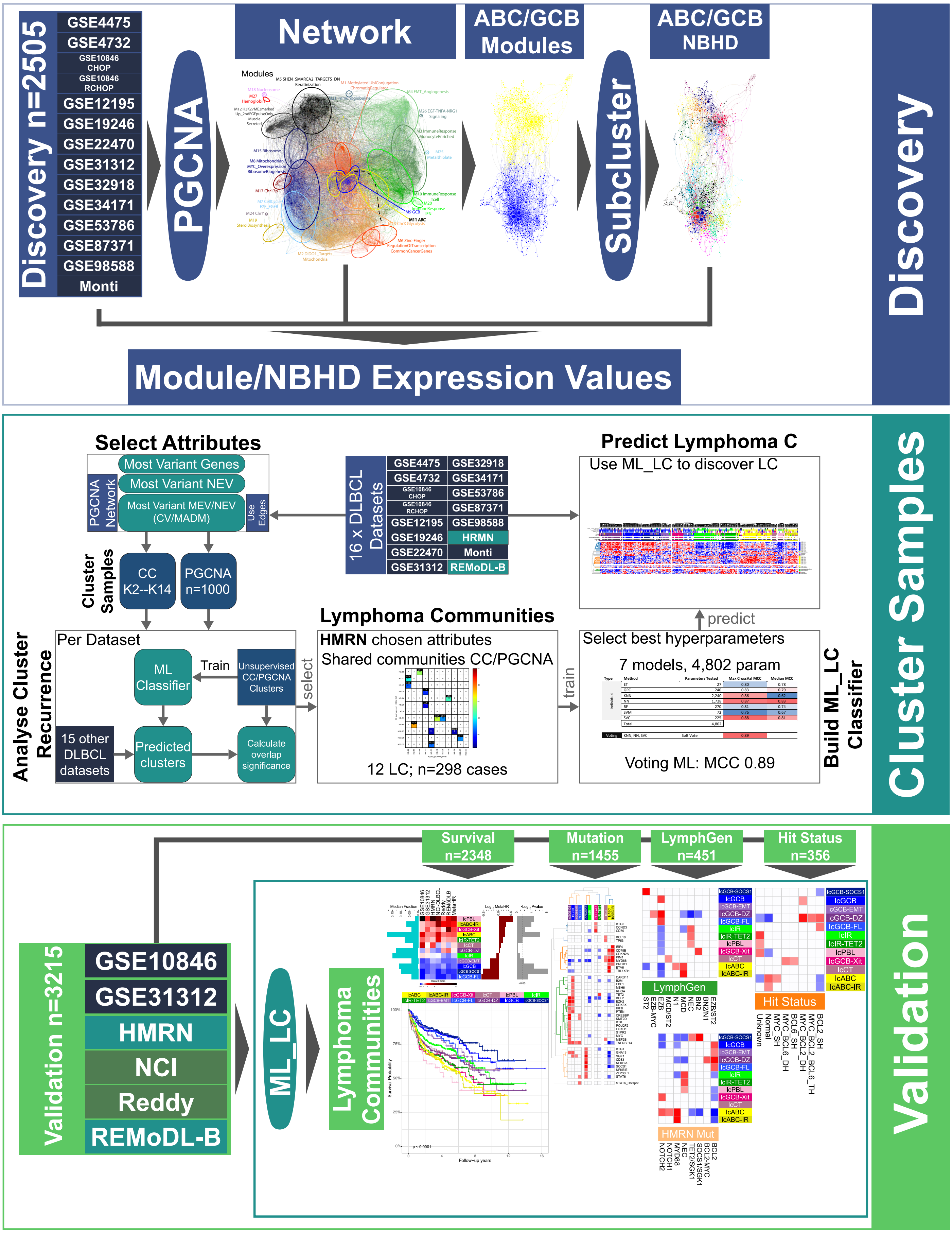


**Extended Data Figure 1. Graphical outline of the methodological steps.**
(**top**) Discovery of network. 14 DLBCL datasets covering 2,505 samples were used to generate a network using PGCNA, the resultant network consisted of 28 modules (Figure 1 and https://mcare.link/DLBCL). All modules were subclustered into neighbourhoods (NBHDs), the ABC/GCB neighbourhoods are shown in Figure 2. The most informative genes per module/neighbourhood were used to generate Module/Neighbourhood Expression Values (MEV/NEV). (**middle**) clustering of samples based on the PGCNA network (see **Supplemental Methods**). Potential attribute sets (gene/MEV/NEVs) are used to cluster 16 DBLCL datasets with Consensus Clustering (CC) and PGCNA approaches. The attribute sets are selected by analysing which provides the most significant cluster rediscovery across the 16 DLBCL datasets. The CC/PGCNA clustering of the HMRN dataset were combined by selecting the shared communities, giving rise to 12 Lymphoma Communities (LC) covering 298 cases. The LC were used to select best ML model hyperparameters, building a final voting classifier (ML_LC) with a MCC=0.89. ML_LC was used to classify 6 DLBCL dataset for downstream validation. (**bottom**) Validation in further held-out datasets. The 12 LC were recovered in 2 additional datasets using the ML_LC tool. The samples from the 6 datasets were used for a combined survival analysis (R-CHOP only, Figure 6) and mutational analysis (Figure 5 and Extended Data Figure 5). Within the HMRN data a comparison to LymphGen/Hit Status (Figure 5) was carried out.
