## Supplemental Figure 2 for "Resolving heterogeneity in Diffuse Large B-cell Lymphoma using a comprehensive modular expression map"

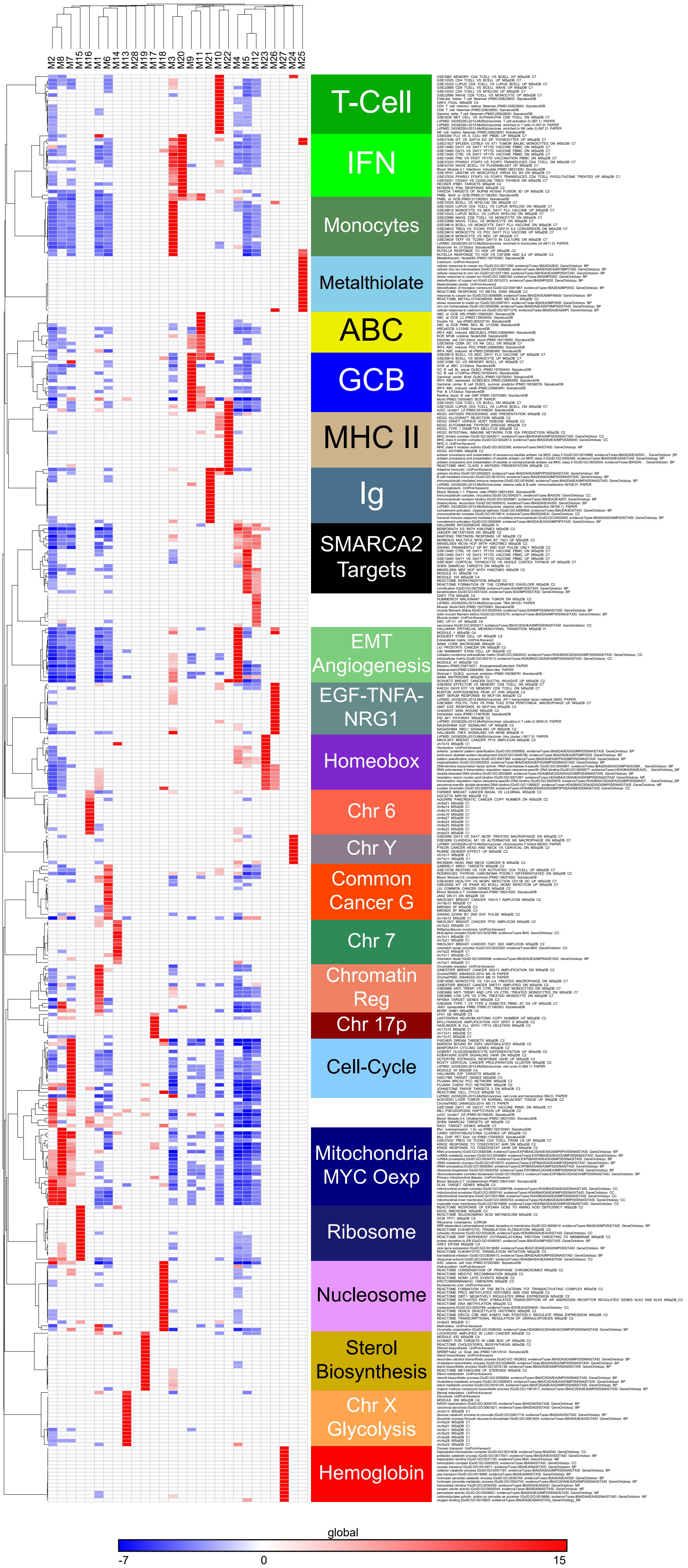


**Extended Data Figure 2. Accompanies Figure 1b. PGCNA resolves distinctive signature and ontology enrichments across network modules**
High resolution version of Figure 1b. Separation of module signature and ontology associations is illustrated as a heatmap (filtered FDR <0.05 and ≥ 5 and ≤ 1000 genes; top 15 most significant signatures per module). Significant enrichment or depletion illustrated on red/blue scale, x-axis (modules) and y-axis (signatures). Hierarchical clustering according to gene signature enrichment. See Supplemental Table 2 for information on all enriched signatures.
