## Supplemental Figure 3 for "Resolving heterogeneity in Diffuse Large B-cell Lymphoma using a comprehensive modular expression map"

**a**

| Module | Description | Modularity | NNum | NSizes |
|---|---|---|---|---|
| M1 | Methylated UblConjugation ChromatinRegulator | 0.652 | 15 | [217, 209, 188, 185, 162, 146, 131, 122, 107, 95, 62, 54, 48, 39, 30] |
| M2 | DIDO1_Targets Mitochondria | 0.638 | 15 | [205, 177, 170, 150, 122, 116, 107, 107, 102, 100, 89, 82, 74, 68, 59] |
| M3 | ImmuneResponse MonocyteEnriched | 0.645 | 13 | [227, 195, 170, 138, 133, 130, 126, 118, 90, 87, 81, 77, 67] |
| M4 | EMT_Angiogenesis | 0.628 | 14 | [228, 212, 171, 168, 163, 156, 113, 95, 66, 57, 51, 24, 21, 5] |
| M5 | SHEN_SMARCA2_TARGETS_DN Keratinization | 0.603 | 15 | [185, 181, 171, 166, 135, 134, 131, 120, 96, 67, 35, 35, 25, 18, 4] |
| M6 | Zinc-Finger RegulationOfTranscription CommonCancerGenes | 0.695 | 14 | [149, 149, 135, 126, 119, 118, 88, 82, 80, 79, 40, 38, 27, 26] |
| M7 | CellCycle E2F_EGFR | 0.649 | 15 | [129, 125, 124, 113, 100, 79, 72, 68, 62, 58, 46, 44, 43, 18, 13] |
| M8 | Mitochondrian MYC_Overexpression RibosomeBiogenesis | 0.667 | 15 | [113, 106, 91, 83, 82, 80, 75, 61, 61, 59, 58, 53, 51, 50, 48] |
| M9 | GCB | 0.649 | 16 | [109, 105, 79, 74, 73, 66, 65, 59, 58, 55, 44, 39, 34, 33, 26, 13] |
| M10 | ImmuneResponse Tcell | 0.623 | 14 | [125, 116, 110, 107, 81, 72, 67, 62, 39, 32, 26, 20, 15, 3] |
| M11 | ABC | 0.698 | 17 | [98, 95, 90, 75, 74, 74, 50, 47, 39, 29, 24, 21, 18, 15, 11, 4, 4] |
| M12 | H3K27ME3marked Up_2ndEGFpulseOnly Muscle Secreted | 0.538 | 11 | [146, 141, 138, 137, 86, 43, 18, 14, 10, 8, 3] |
| M13 | ChrX Glycolysis | 0.684 | 12 | [54, 35, 33, 30, 30, 26, 20, 18, 15, 12, 12, 11] |
| M14 | Chr7 | 0.702 | 9 | [31, 27, 26, 26, 23, 19, 18, 5, 4] |
| M15 | Ribosome | 0.596 | 9 | [26, 24, 19, 19, 18, 16, 15, 15, 9] |
| M16 | Chr6 | 0.636 | 9 | [25, 25, 23, 20, 17, 14, 12, 7, 7] |
| M17 | Chr17p | 0.574 | 6 | [19, 18, 15, 14, 12, 9] |
| M18 | Nucleosome | 0.496 | 4 | [16, 16, 12, 6] |
| M19 | SterolBiosynthesis | 0.529 | 4 | [13, 13, 7, 7] |
| M20 | ImmuneResponse IFN | 0.506 | 4 | [12, 11, 9, 7] |
| M21 | Immunoglobulins | 0.567 | 4 | [11, 10, 8, 6] |
| M22 | MHC II | 0.394 | 4 | [11, 8, 7, 3] |
| M23 | Homeobox | 0.364 | 2 | [7, 5] |
| M24 | ChrY Enriched | 0.202 | 3 | [4, 4, 4] |
| M25 | Metalthiolate | 0.098 | 2 | [6, 3] |
| M26 | EGF-TNFA-NRG1_Signaling | 0.098 | 2 | [4, 4] |
| M27 | Hemoglobin | 0.045 | 2 | [3, 3] |
| M28 | Novel1 | 0.100 | 2 | [3, 3] |

**b**

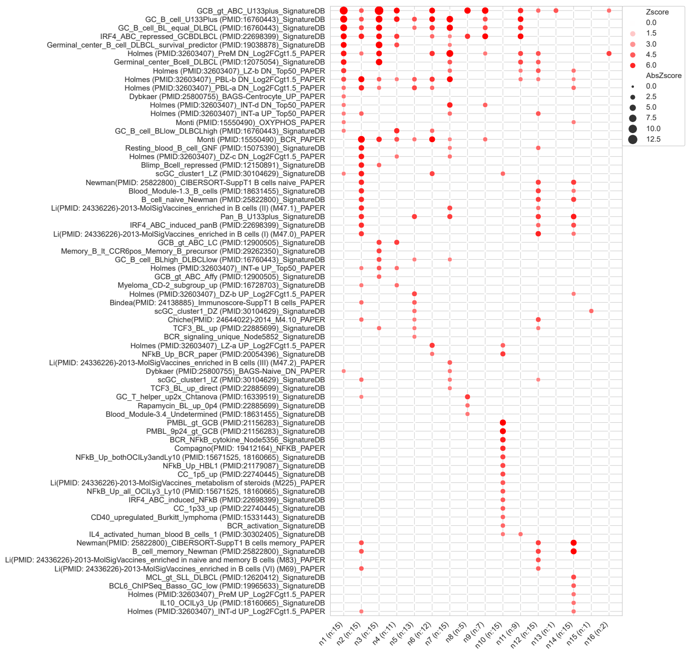


| Neighbourhood | GSE | Type |
|---|---|---|
| 1 | LZ/Centrocyte_Up LZb_DN PBL-a/b_DN | LZ |
| 2 | Int-a/e/d_UP PBL-a/b_DN DZ-C DN | Int |
| 3 | Int-e_UP Memory_B_lt_CCR6posMemB PreM_DN | Int |
| 4 | BL-Low_DLBCL-High Int-e_UP | Int |
| 5 | DZ-b_UP PBL-a/b_DN | DZ |
| 6 | LZ-a_UP PBL-a/b_DN PreM_DN NFKB | LZ-NFKB |
| 7 | PreM_DN BCL6_ChIPseq Int-a/b_UP BL-High_DLBCL_Low | Int |
| 8 | GCB | |
| 9 | BL_equal_DLBCL Int-d_DN PreM_DN | |
| 10 | PMBL_gt_GCB NFKB LZ-a_UP | LZ-NFKB |
| 11 | BL_equal_DLBCL PreM_DN PBL-b_DN LZ-b_DN | |
| 12 | Int-a_UP LZ-a/b_DN PBL-b/DN | Int |
| 13 | TP53 Signalling Regulated-Necrosis | TP53 |
| 14 | DZ-b_UP PBL-a/b_DN PreM_UP LZ-b_DN | DZ |
| 15 | scGC_cluster1_DZ | DZ |
| 16 | PreM_DN | |

DAC classifier genes

| Class | Gene | Module | ModuleNeighbourhood |
|---|---|---|---|
| GCB | BCL6 | 9 | 3 |
| GCB | MME | 9 | 3 |
| GCB | SERPINA9 | 9 | 3 |
| GCB | DENND3 | 9 | 4 |
| GCB | ITPKB | 9 | 4 |
| GCB | LMO2 | 9 | 4 |
| GCB | NEK6 | 9 | 4 |
| GCB | IRAG2 (LRMP) | 9 | 11 |

**c**

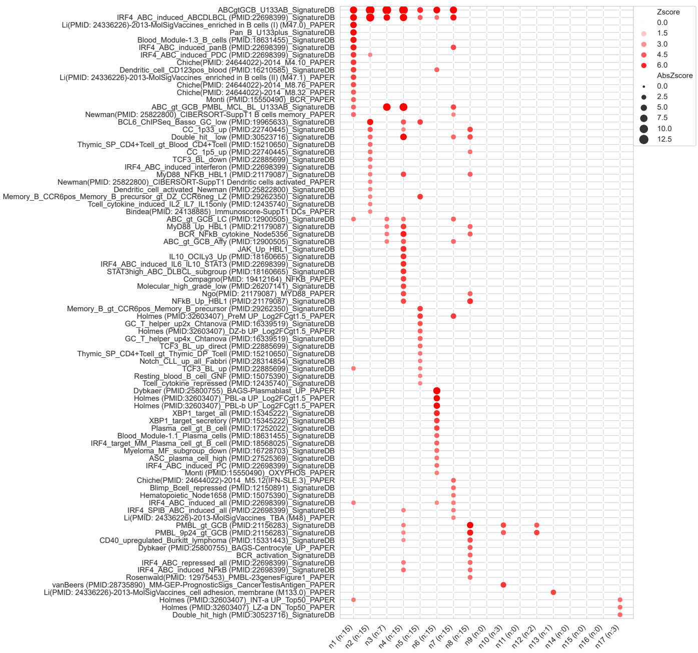


| Neighbourhood | GSE | Type |
|---|---|---|
| 1 | Bcell_Activation DendriticCell Int-a_UP Int-e_DN | Int |
| 2 | Centrocyte_UP MYD88_NFKB DendriticActivated | LZ |
| 3 | MYD88_NFKB | |
| 4 | BCR_NFkB JAK_Up STAT3high MYD88 MHG_Low | |
| 5 | Memory_B_gt_CCR6posMemB DZ-b_UP TCF3_BL_UP Pre | PreM |
| 6 | Plasmablast PBL-a/b_UP XBP1_Targets | PBL |
| 7 | PreM_UP BLIMP1_Repressed | PreM |
| 8 | NFKB Centrocyte_Up MYD88 PMBL | LZ; PMBL |
| 9 | Chr18q21-22 | ChromRegion |
| 10 | CancerTestisAntigen PMBL_gt_GCB | PMBL |
| 11 | Novel1 | |
| 12 | PMBL_gt_GCB | PMBL |
| 13 | CellAdhesion | |
| 14 | Novel2 | |
| 15 | Chr18q21 | ChromRegion |
| 16 | Novel3 | |
| 17 | Chr6q21 | ChromRegion |

DAC classifier genes

| Class | Gene | Module | ModuleNeighbourhood |
|---|---|---|---|
| ABC | BLNK | 11 | 1 |
| ABC | FOXP1 | 11 | 1 |
| ABC | CCND2 | 11 | 2 |
| ABC | ETV6 | 11 | 2 |
| ABC | BMF | 11 | 3 |
| ABC | IRF4 | 11 | 3 |
| ABC | PIM1 | 11 | 3 |
| ABC | ENTPD1 | 11 | 4 |
| ABC | FUT8 | 11 | 4 |
| ABC | PTPN1 | 11 | 4 |
| ABC | IL16 | 11 | 7 |
| ABC | SH3BP5 | 11 | 7 |

**Extended Data Figure 3. Accompanies Figure 2. Neighbourhood analysis provides fine grained assessment of gene correlations**
Neighbourhood Analysis **(a)** Tabulated summary of neighbourhood analysis for all network modules. Ordered by module number, showing best modularity score (5,000 leidenalg cluster runs) with corresponding number of neighbourhoods and gene number within each neighbourhood. **(b)** and **(c)** Detailed version of Figure 2 (b) and (c) showing separation of neighbourhood signature and ontology associations as a bubble plot for M9_GCB (b) and M11_ABC (c) (filtered FDR <0.05 and ≥ 5 and ≤ 4000 genes; top 15 most significant signatures per neighbourhood). Significant enrichment illustrated by colour and bubble size, x-axis (neighbourhoods) and y-axis (signatures). The number of selected signatures per neighbourhood is shown in parenthesis after each neighbourhood name. For all signatures see Supplemental Table 4. Below each bubble plot is a table with signature summaries for each neighbourhood and a table showing which neighbourhood the cell-of-origin classifier genes belong to.
