## Supplemental Figure 4 for "Resolving heterogeneity in Diffuse Large B-cell Lymphoma using a comprehensive modular expression map"

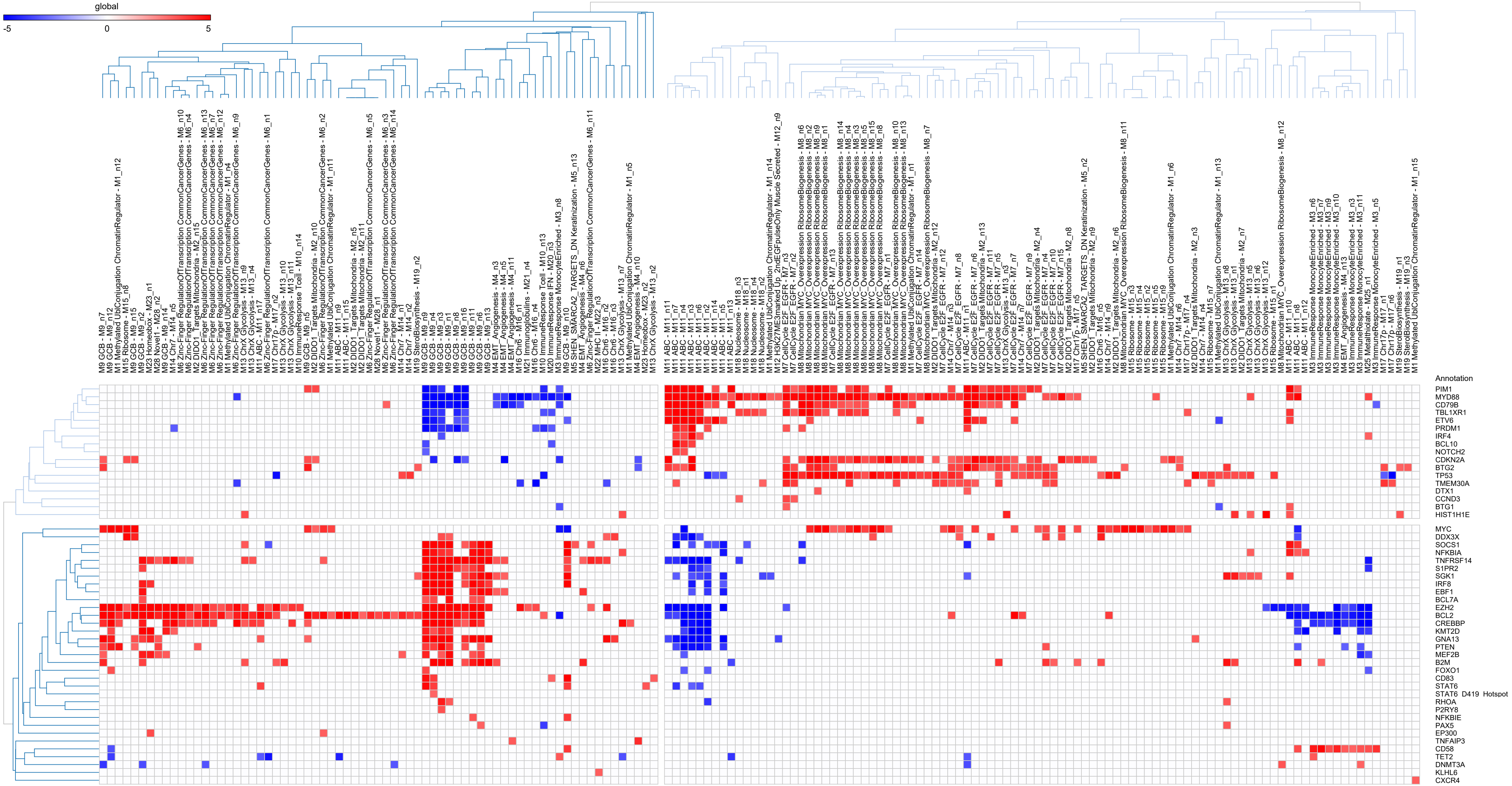


**Extended Data Figure 4. Relationship between NEVs and mutations**

Relates to Figure 3. Association of Neighbourhood Expression Values (NEV) with mutations. Shown are combined significance (Stouffer method) of p-value based on point-biserial correlations between binary mutation status and MEV/NEVs across 3 datasets (HMRN, Reddy & REMoDLB). Only mutations with p-value < 0.001, occurring in ≥ 2 datasets were retained, Z-scores for p-values > 0.001 were set to 0. Significance is shown as z-scores on a blue (significant depletion) to red (significant enrichment) scale (-5 to +5).
