## Supplemental Figure 5 for "Resolving heterogeneity in Diffuse Large B-cell Lymphoma using a comprehensive modular expression map"

GSE10846 R-CHOP

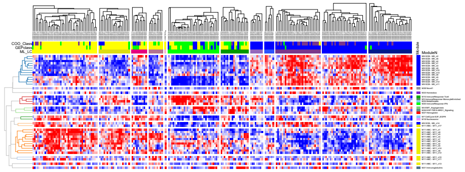

GSE31312

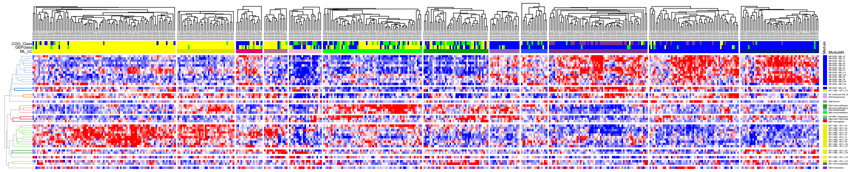

HMRN

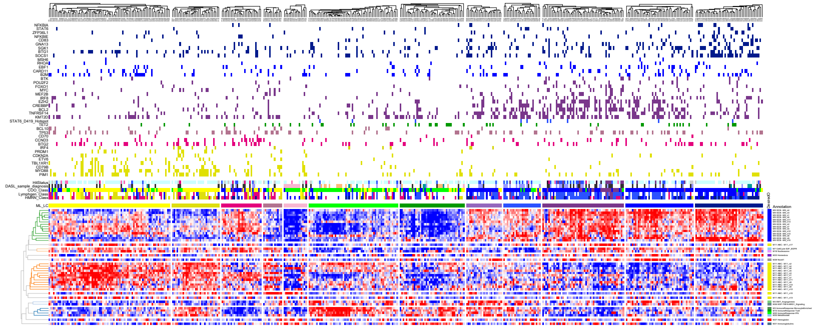

NCI-DLBCL

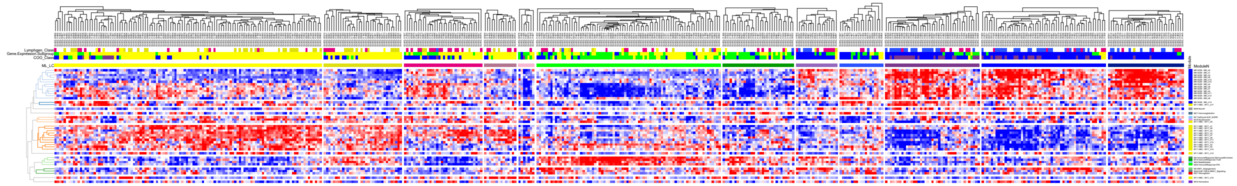

Reddy

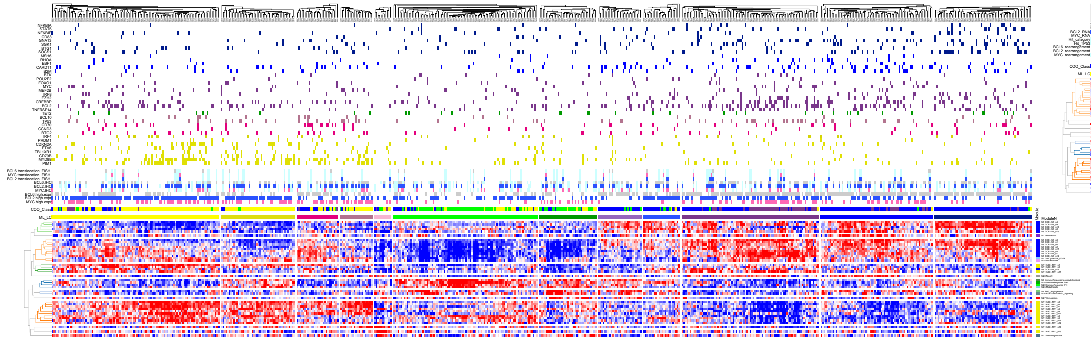

REMoDLB (All Data)

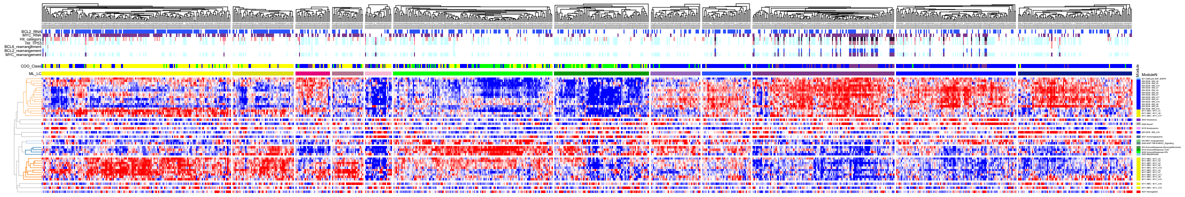

REMoDLB (With Mutations)

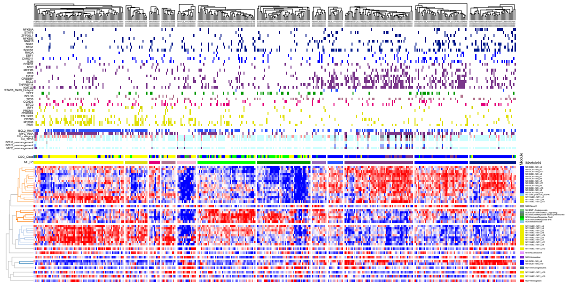

**Extended Data Figure 5. Heatmaps for 6 datasets used for survival analysis**
Relates to figure 4—8. Shows the expression levels of Module Expression Values (MEVs) and Neighbourhood Expression Values (NEVs; for M9:GCB and M11:ABC) for the 6 datasets used for survival analysis. The dataset are grouped by the 12 Lymphoma Communities assigned by the machine learning tool (ML_LC) which was trained on a subset of the HMRN data (see supplemental methods). Heatmaps show cases hierarchically clustered within each ML_LC group. MEV and NEV are shown on a blue (low) to red (high) z-score colour scale. Modules and neighbourhoods are separated across the y-axis and colour coded by their module. Meta-data is provided above the heatmap including individual case level mutation data as well as COO and MHG class, LymphGen and HMRN mutational classifications, rearrangement hit status for MYC, BCL2 and BCL6 and morphological classification, see legend in bottom right corner for details.

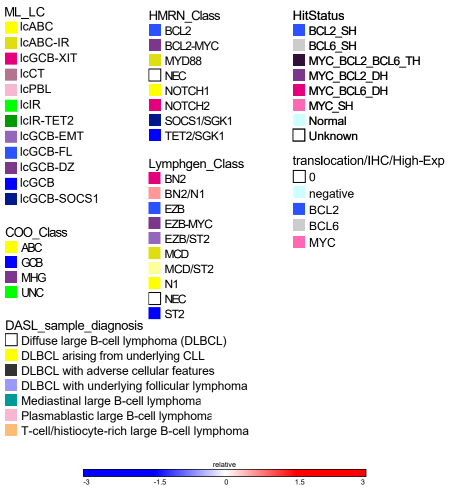
