## Supplemental Figure 6 for "Resolving heterogeneity in Diffuse Large B-cell Lymphoma using a comprehensive modular expression map"

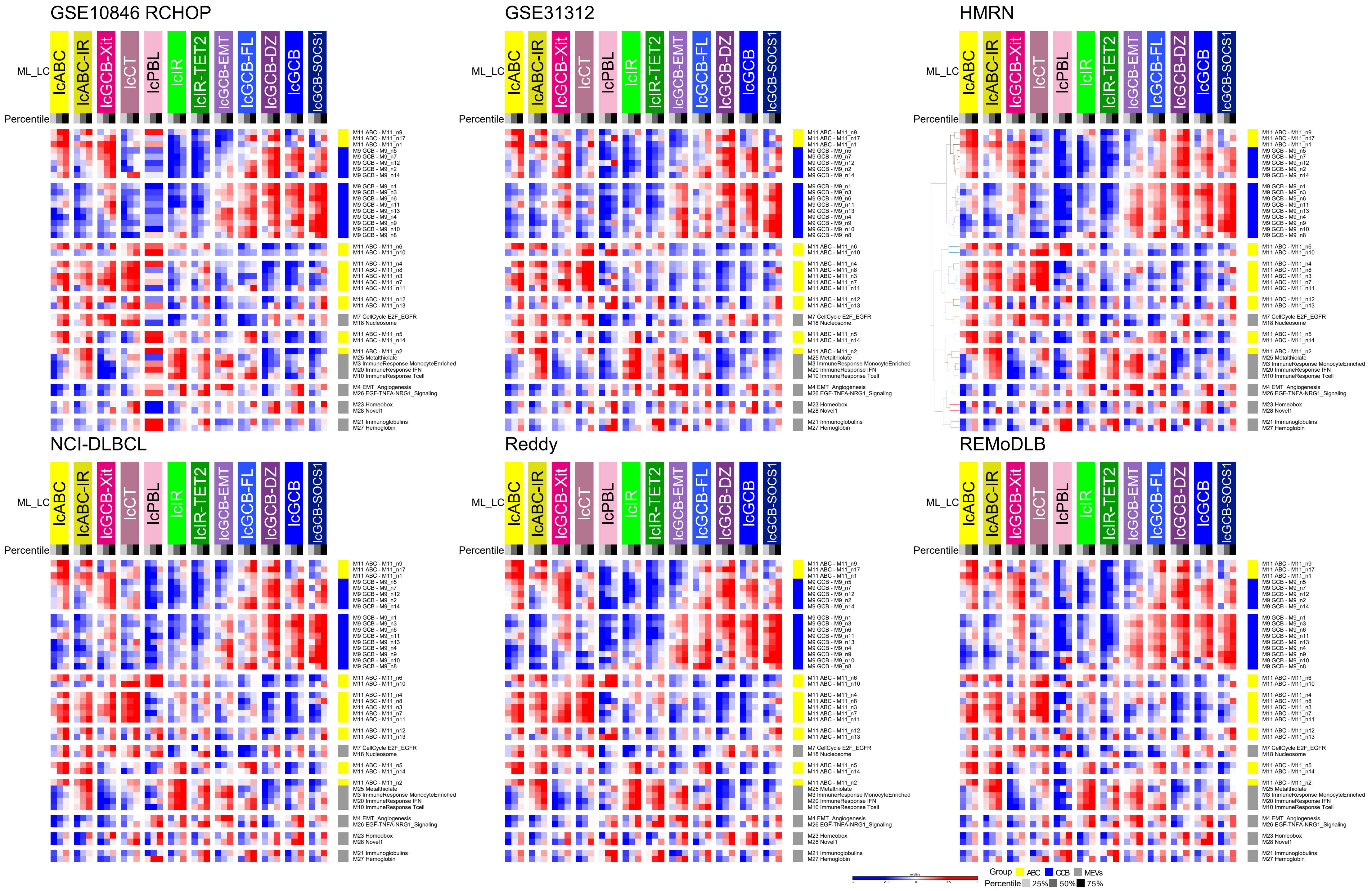


**Extended Data Figure 6. Accompanies Figure 4. Lymphoma Communities (LC) IQR for each dataset.**

Shows the patterns of MEV and NEV (y-axis) expression across the 12 LC (x-axis; ML_LC) in the GSE10846 (RCHOP), GSE31312, HMRN, NCI-DLBCL, Reddy and REMoDL-B datasets. Expression is illustrated on z-score blue to red scale as an interquartile range plot, with dendrogram of MEV/NEV hierarchical clustering shown on left (HMRN only). All datasets were forced to have the same order as the hierarchically clustered HMRN dataset to aid comparison.
