## Supplemental Figure 7 for "Resolving heterogeneity in Diffuse Large B-cell Lymphoma using a comprehensive modular expression map"

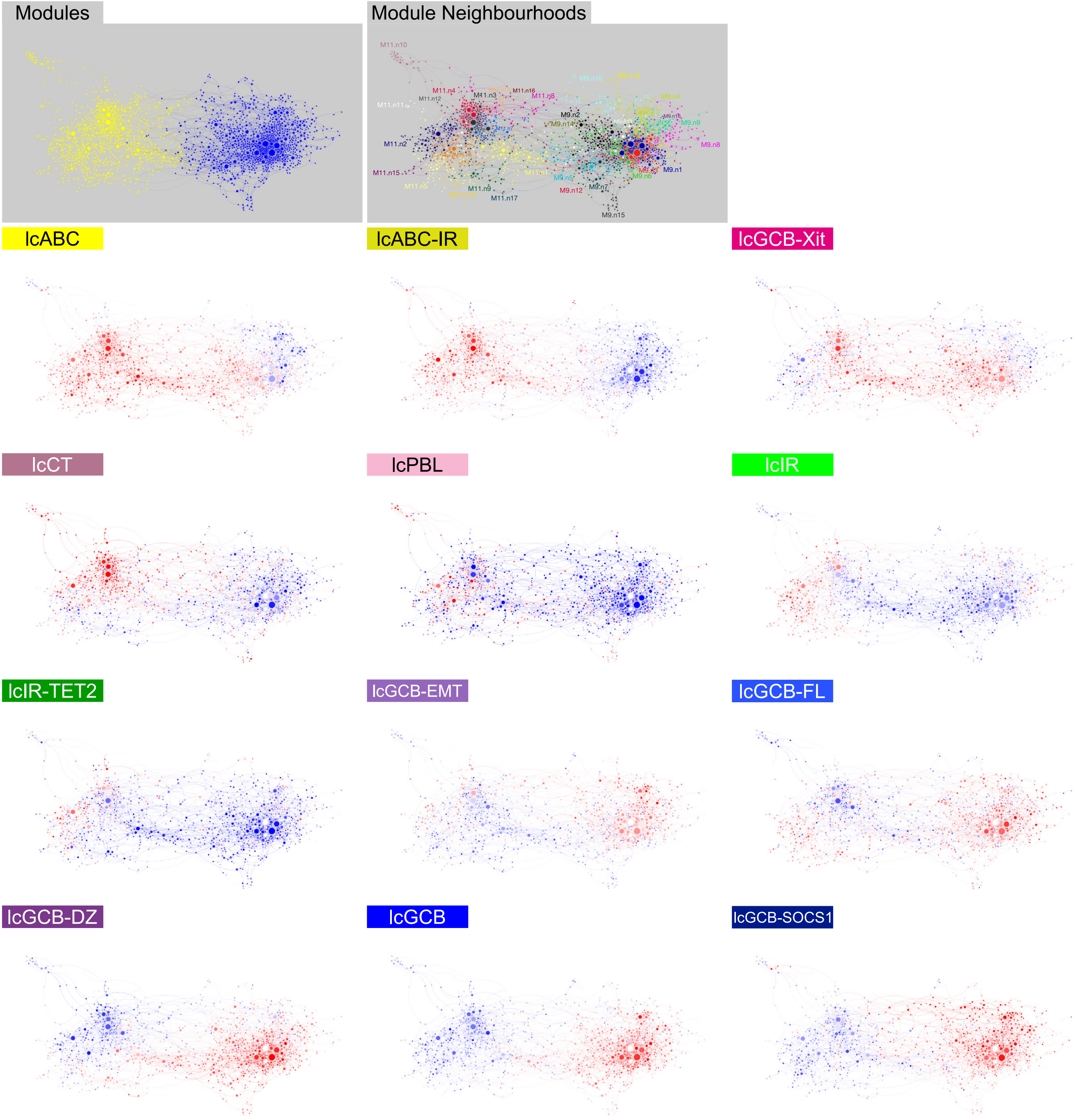


**Extended Data Figure 7. Accompanies Figure 4. Lymphoma Communities (LC) median expression values displayed across GCB/ABC neighbourhood network visualization**

Relationship of median gene expression for all LC is displayed in the context of the ABC/GCB neighbourhood network. Median gene expression values for all genes (in HMRN dataset) in the network are displayed for each LC as blue to red z-score scale.
