## Supplemental Figure 8 for "Resolving heterogeneity in Diffuse Large B-cell Lymphoma using a comprehensive modular expression map"

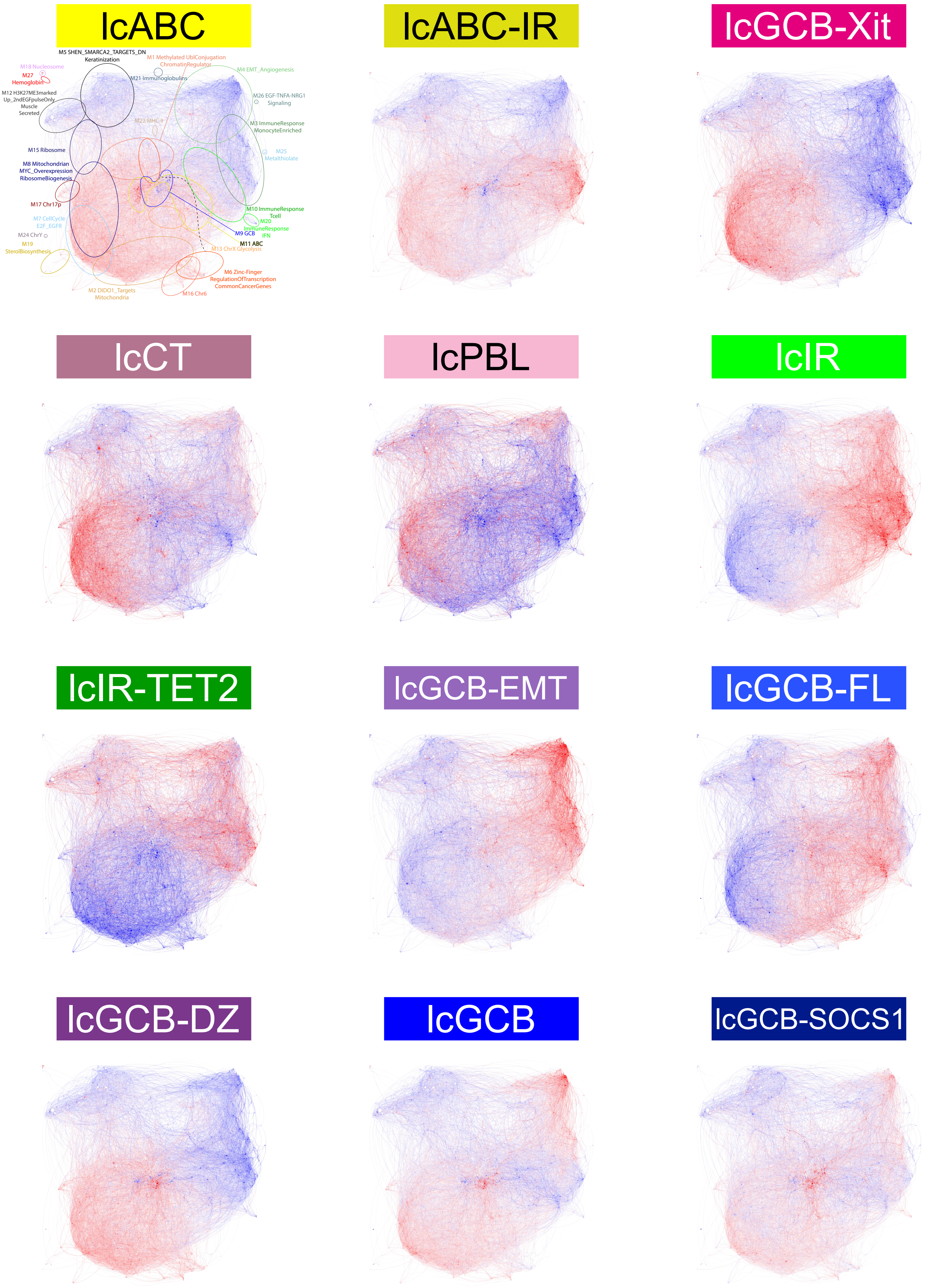


**Extended Data Figure 8. Accompanies Figure 4. Lymphoma Communities (LC) median expression values displayed across DLBCL network visualization.**

Relationship of all gene expression across all LC is displayed in the context of the overall DLBCL network as in Figure 1a. Median gene expression values for all genes (in HMRN dataset) in the network are displayed for each LC as blue to red z-score scale.
