## Supplemental Figure 10 for "Resolving heterogeneity in Diffuse Large B-cell Lymphoma using a comprehensive modular expression map"

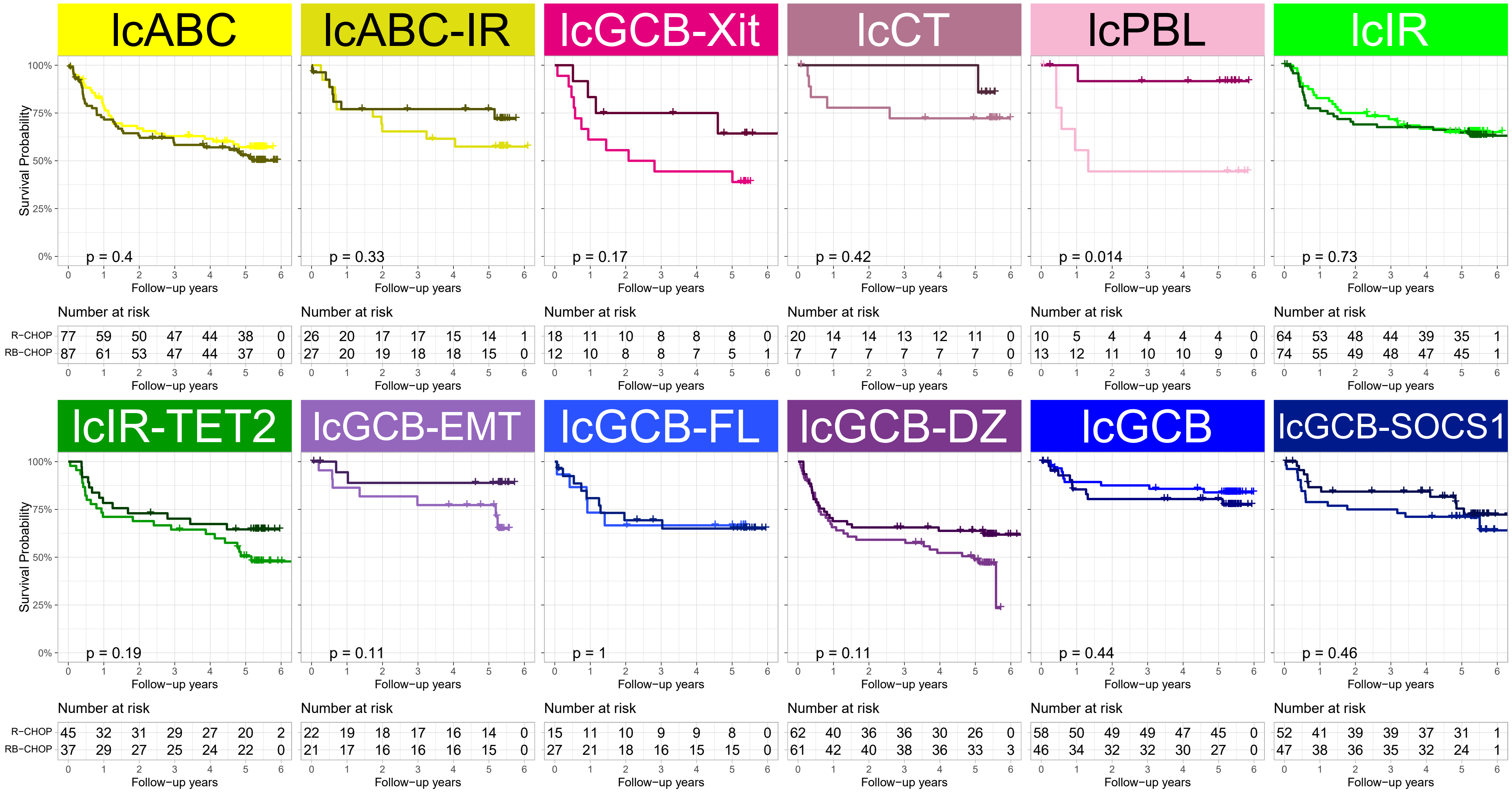


**Extended Data Figure 10. Progression Free Survival across the Lymphoma Communities split by treatment.**
Shows Kaplan-Meier plots of survival for Lymphoma Communities, each plot comparing the treatments: R-CHOP (lighter-line) and RB-CHOP (darker-line) along with p-value from log-rank test.
