## Supplemental Methods for "Resolving heterogeneity in Diffuse Large B-cell Lymphoma using a comprehensive modular expression map"

### Contents


Gene expression analysis
- Expression datasets
  - Network generation
  - Clustering Samples
  - Validation
- Normalization and re-annotation of data

PGCNA network
- Background
- PGCNA network generation
- Network meta-data
- Network visualisation

Clustering samples
- Neighbourhoods
- Module expression values (MEV)
- Selection of genes/MEV/NEV
  - Ignoring edges
  - Using edges
- Clustering approaches
- Assess cluster recurrence using machine learning
- Lymphoma Communities (LC)

Machine learning
- Input data
- Training and validation datasets
- Model selection
- Classification of datasets

Statistical analyses
- Gene signature data and enrichment analysis

Survival analysis
- Gene/LC level meta-analysis

Mutation analysis
- MEV/NEV analysis
- LC analysis
- Mutation meta-analysis

LC differential gene expression

Data visualisations
- LC visualisations
- Heatmaps
- Violin plots

Software and data
Data processing
References

### Gene expression analysis

#### Expression datasets

The datasets used in the paper are outlined in Table 1 (and ST1), see EDF1 for overview of analysis described in the supplemental methods.

##### Network generation

The Gene Expression Omnibus (GEO) was searched for DLBCL data sets with > 50 patient samples (search date: 2019.01.09), providing a representative set of data. For network generation, 13 DLBCL gene expression datasets were downloaded from the Gene Expression Omnibus (GSE10846 was split into CHOP/R-CHOP treated).[3] In total these datasets contain 2,505 DLBCL samples (see PGCNA network)

##### Clustering Samples

Once the network had been created and the modules/neighbourhoods defined (see PGCNA network generation and Neighbourhoods) then 16 dataset (addition of HMRN/REMoDLB; see ST1) were used to select attributes and cluster samples into Lymphoma Communities (LC see Clustering samples), these formed the training classifications for machine learning (ML, see Machine learning), and were used for downstream analysis within 6 datasets (including 2 additional validation datasets; see ST1).

##### Validation

2 datasets were held-out for validation of the ML assigned LC groups. These datasets covered an additional 1,105 DLBCL cases.

#### Normalization and re-annotation of data

For the RNA-seq datasets (NCI, Reddy) the count data was processed using DESeq2 v1.22.2 with VST normalised data used for downstream analysis.[1] For each data set the probes were re-annotated using the MyGene.info (http://mygene.info) API using all available references (e.g. NCBI Entrez, Ensembl etc.) and any ambiguous mappings manually assigned[2]. Each data set was quantile normalized using the Python qnorm package and the probes for each gene merged by taking the median value for probe sets with a Pearson correlation ≥0.2 and the maximum value for those with a correlation <0.2.

### PGCNA network

#### Background

For details and validation of the Parsimonious Gene Correlation Network Analysis (PGCNA) approach see our other work.[4] Here a brief description of the method will be given. After informative genes are selected they are used to calculate Spearman's rank correlations for all gene pairs using the Python scipy.stats package. For each gene (row) in a correlation matrix only the 3 most correlated edges per gene are retained. The resulting matrix $M$, with entries written as $M = (m_{ij})$ is made symmetrical by setting $m_{ij} = m_{ji}$ for all indices $i$ and $j$ so that $M = M^T$ (its transpose). The correlation matrices are clustered using a community detection algorithm (Leidenalg v0.8.3) and the best (judged by modularity score) used for downstream analysis. [5]

#### PGCNA network generation

PGCNA2 (https://github.com/medmaca/PGCNA/tree/master/PGCNA2) was run with the following command:

| Type | Source | Author | PubMed ID | Publish Year | No. samples in study | hosen Sampl | Survival Data | Detail.sample |
|---|---|---|---|---|---|---|---|---|
| DLBCL Network-Generation | GSE4475 | Hummel M. | 16760442 | 2006 | 221 | 129 | Yes | 44mBL, 48intermediate, 129non-mBL |
| | GSE4732 | Dave SS. | 16760443 | 2006 | 303 | 66 | Yes | 33BL, 22ABC, 24GCB, 20PMBL |
| | GSE10846_CHOP | Lenz Z. | 19038878 | 2008 | 420 | 181 | Yes | DLBCL (233RCHOP, 181CHOP) |
| | GSE10846_RCHOP | Lenz Z. | 19038878 | 2008 | 420 | 233 | Yes | DLBCL (233RCHOP, 181CHOP) |
| | GSE12195 | Pasqualucci I | 19412164 | 2009 | 136 | 73 | | 73 DLBCL, 38 FL, 5 cell lines |
| | GSE19246_FF | Williams PM | 20688907 | 2010 | 177 | 59 | | 59 DLBCL_FF |
| | GSE22470 | Salaverria I | 21487109 | 2011 | 271 | 271 | | DLBCL |
| | GSE31312 | Frei E. | 23775435 | 2011 | 498 | 498 | Yes | DLBCL FFPE |
| | GSE32918 | Care M | 24875472 | 2013 | 249 | 171 | Yes | 171 DLBCL FFPE |
| | GSE34171 | Chapuy B | 22975378 | 2012 | 169 | 169 | Yes | DLBCL (78 Affy 133A+B, 91 Affy133+2) |
| | GSE53786 | Scott DW | 24398326 | 2014 | 119 | 119 | Yes | DLBCL 119 Retrospetive FFPE samples; 45 overlap with GSE4732 |
| | GSE87371 | Dubois S | 27923841 | 2017 | 223 | 223 | Yes | DLBCL LYSA trials |
| | GSE98588 | Chapuy B | 29713087 | 2018 | 137 | 137 | Yes | DLBCL |
| | Monti | Monti S. | 15550490 | 2005 | 176 | 176 | Yes | DLBCL |
| | | | | | Total | 2505 | | |
| + For Sample Clustering | HMRN (GSE181063) | | | | 1310 | 451 | Yes | DLBCL FFPE; 451 with expression and mutation information |
| | REMoDLB (GSE117556) | Sha | 30523719 | 2018 | 928 | 928 | Yes | 928 DLBCL FFPE (REMoDLB trial) |
| Validation | NCI (NCICCR-DLBCL) | Schmitz | 29641966 | 2018 | 481 | 481 | Yes | DLBCL; 234 with survival information |
| | Reddy (EGAS00001002606) | Reddy | 28985567 | 2017 | 624 | 624 | Yes | DLBCL |
| | | | | | Total | 2484 | | |

Table 1: datasets used for network generation and validation

```
pgcna2 -w . -d /networkData -m '#FileInfo.txt' -s "\t" -f 0.7 -g 0.5 -e 3 -n 1000 -b 100 -r 4 --keepBigF –singleCorr
```

Retaining the top 70% most variant genes present in at least 50% of the datasets, carrying out 1000 Leidenalg clusterings of the data and selecting the best using Scaled cluster enrichment scores (see original PGCNA methods for details). [4] This resulted in a network containing 16,054 genes (44,730 edges) that was split into 28 modules (see ST1).

#### Network meta-data

Several additional features were calculated for the network genes across the data sets used to generate the correlation network. For each gene the median percentile expression was calculated across all data sets, its dispersion across datasets calculated as the median absolute deviation (MAD) and its dispersion within datasets (i.e. across patients) calculated as the median quantile coefficient of dispersion (QCOD).

#### Network visualisation

The PGCNA edges/node files were uploaded into the Gephi package (version 0.9.2)[6]. Node size was set by degree and the network layout generated using the ForceAtlas2 approach, and interactive HTML5 web visualizations exported using the sigma.js library (https://github.com/oxfordinternetinstitute/gephi-plugins/tree/sigmaexporter-plugin). Interactive networks are available at https://mcare.link/DLBCL.

### Clustering samples

The PGCNA network contains modules consisting of genes with similar expression patterns. We wanted to use this information to stratify the datasets into communities of samples that expressed similar modules, and thus had similar underlying biology (see EDF1: Cluster Samples).

Before clustering the data, we first split each module into submodules/neighbourhoods (Neighbourhoods) and then collapsed the genes within these to single values (MEV/NEV; Module expression values). We selected the most informative genes/MEV/NEVs using a number of different approaches (Selection of genes/MEV/NEV), and then used these sets to cluster the DLBCL datasets using two different approaches PGCNA/CC (Clustering approaches). The cluster results were used to explore how recurrently discoverable the DLBCL communities were (Assess cluster recurrence using machine learning) allowing the selection of the most informative attributes for clustering. The best results from this for the HMRN dataset were then combined between PGCNA/CC to generate ensemble Lymphoma Communities (Lymphoma Communities (LC)). Finally, these were used to train a machine learning tool to recover the DLBCL Lymphoma Communities in every dataset (Machine learning).

#### Neighbourhoods

Each of the 28 modules was sub-clustered to create neighbourhoods. The existing PGCNA edge file was split at the module level and then clustered 5,000 times using Leidenalg. The best clustering (based on modularity score) for each module was retained. Multiple different runs converged onto the same answer. The neighbourhoods are show in Table 2.

| Module | Modularity | NNum | NSizes |
|---|---|---|---|
| M1 | 0.652 | 15 | [217, 209, 188, 185, 162, 146, 131, 122, 107, 95, 62, 54, 48, 39, 30] |
| M2 | 0.638 | 15 | [205, 177, 170, 150, 122, 116, 107, 107, 102, 100, 89, 82, 74, 68, 59] |
| M3 | 0.645 | 13 | [227, 195, 170, 138, 133, 130, 126, 118, 90, 87, 81, 77, 67] |
| M4 | 0.628 | 14 | [228, 212, 171, 168, 163, 156, 113, 95, 66, 57, 51, 24, 21, 5] |
| M5 | 0.603 | 15 | [185, 181, 171, 166, 135, 134, 131, 120, 96, 67, 35, 35, 25, 18, 4] |
| M6 | 0.695 | 14 | [149, 149, 135, 126, 119, 118, 88, 82, 80, 79, 40, 38, 27, 26] |
| M7 | 0.649 | 15 | [129, 125, 124, 113, 100, 79, 72, 68, 62, 58, 46, 44, 43, 18, 13] |
| M8 | 0.667 | 15 | [113, 106, 91, 83, 82, 80, 75, 61, 61, 59, 58, 53, 51, 50, 48] |
| **M9** | 0.649 | 16 | [109, 105, 79, 74, 73, 66, 65, 59, 58, 55, 44, 39, 34, 33, 26, 13] |
| M10 | 0.623 | 14 | [125, 116, 110, 107, 81, 72, 67, 62, 39, 32, 26, 20, 15, 3] |
| **M11** | 0.698 | 17 | [98, 95, 90, 75, 74, 74, 50, 47, 39, 29, 24, 21, 18, 15, 11, 4, 4] |
| M12 | 0.538 | 11 | [146, 141, 138, 137, 86, 43, 18, 14, 10, 8, 3] |

| | | | |
|---|---|---|---|
| M13 | 0.684 | 12 | [54, 35, 33, 30, 30, 26, 20, 18, 15, 12, 12, 11] |
| M14 | 0.702 | 9 | [31, 27, 26, 26, 23, 19, 18, 5, 4] |
| M15 | 0.596 | 9 | [26, 24, 19, 19, 18, 16, 15, 15, 9] |
| M16 | 0.636 | 9 | [25, 25, 23, 20, 17, 14, 12, 7, 7] |
| M17 | 0.574 | 6 | [19, 18, 15, 14, 12, 9] |
| M18 | 0.496 | 4 | [16, 16, 12, 6] |
| M19 | 0.529 | 4 | [13, 13, 7, 7] |
| M20 | 0.506 | 4 | [12, 11, 9, 7] |
| M21 | 0.567 | 4 | [11, 10, 8, 6] |
| M22 | 0.394 | 4 | [11, 8, 7, 3] |
| M23 | 0.364 | 2 | [7, 5] |
| M24 | 0.202 | 3 | [4, 4, 4] |
| M25 | 0.098 | 2 | [6, 3] |
| M26 | 0.098 | 2 | [4, 4] |
| M27 | 0.045 | 2 | [3, 3] |
| M28 | 0.100 | 2 | [3, 3] |

Table 2: neighbourhood clustering. Showing the best clustering for each module, the modularity score (Modularity) the neighbourhood-number (NNum) and the spread of neighbourhood sizes (NSizes).

#### Module expression values (MEV)

The genes per module/neighbourhood were collapsed down to single values in the following manner:

Within each data set, which vary in available genes, the genes per module/neighbourhood were ranked by gene_strength (sum of genes edges/correlations within its module).

Once the representative genes have been selected, they are converted into a Module Expression Value (MEV) or Neighbourhood Expression Value (NEV) by:

1. Per module/neighbourhood select top 10 genes based on ranks.
2. Per gene, standardize (z-score) the quantile normalized $\log_2$ expression data.
3. Per sample (patient) calculate the median of the 10 z-scores to give a MEV/NEV.

Used in Fig 4, EDF5.

#### Selection of genes/MEV/NEV

The PGCNA network splits the gene expression data into modules/neighbourhoods based on the correlation structure of the genes. To cluster samples, we aim to use these modules/neighbourhoods to split the data into groups with similar modular expression patterns. However, there are various approaches that could be used:

- Per dataset could just select the most variant genes (using Median Absolute Deviation, Coefficient of variation etc.)
    - Depending on number of genes chosen this might miss important features.
    - The chosen genes might differ between datasets making comparisons harder (important if you're trying to find recurrent modules across datasets).
- Alternatively, you could use the PGCNA network neighbourhoods to ensure a fair representation of the different features that the network captures, can do this in two different ways:
    - Use network structure:
        - Select most informative genes per module/neighbourhood based on gene degree.
        - Rank module/neighbourhoods by variance (CV; MADM etc).
        - Select module/neighbourhoods to retain.
    - Only use module/neighbourhoods:
        - Select most variant gene per module/neighbourhood (ignoring network structure).

Each of the approaches discussed below was applied to all 18 datasets (see ST1) to recover sets of genes/MEV/NEV that were informative in as many datasets as possible.

#### Ignoring edges

The first approach (SelectG_MADS) selects the most informative genes per module/neighbourhood without using information on connectivity (i.e., the gene degree):

Using (Median Absolute Deviation)/Median (MADM) to select the 5 most variant genes per module/neighbourhood:

- Per dataset (n=18):
  - Find MADM per gene.
  - Calculate median of MADM across datasets.
- Filter genes:
  - Remove genes present < 14 datasets.
  - Per neighbourhood calculate median MADM of top 5 genes:
    - Remove neighbourhood if < 60$^{th}$ percentile across all median MADM.
- Write out:
  - Per dataset select top 5 highest available ranked genes per retained MEV/NEV.

#### Using edges

The two methods (SelectG_NEV-CV & SelectG_NEV-MADM) use the network structure to inform gene selection. They use either Coefficient of Variation (CV), or (Median Absolute Deviation)/Median (MADM) to filter genes using the following approach (below CV or MADM is used accordingly):

Select most informative MEV/NEVs based on their median CV across a set of datasets.

For each dataset:

- Select the highest ranked genes (based on strength; sum of correlations) for each module/neighbourhood (n=10 genes).
- Calculate the coefficient of variation (CV) for each of these genes.
- Calculate the median of these CV to generate the median CV for that module/neighbourhood.
- Repeat across all modules/neighbourhoods to generate a distribution of median CVs.
- Generate random data (n=1000):
  - Randomly select a set of genes that equals the number of genes spread across all modules/neighbourhoods.
  - Split into groups to size match the actual modules/neighbourhoods.
  - Calculate median CVs per module/neighbourhood.
- Using a cut-off that is 3 standard deviations (> 99.7% rand) above the mean in the random data note which modules/neighbourhoods are “significant”. This aims at finding the MEV/NEVs that have a large amount of variance across the samples.
- Repeat across all datasets.
- Retain all modules/neighbourhoods that are "significant" in 0.5 of the provided datasets (minDS0.5).
- Output this set of modules/neighbourhoods for downstream work. Both at gene level and collapsed to MEV/NEVs.

### Clustering approaches

The genes/MEV/NEV derived from Selection of genes/MEV/NEV were then used to cluster each dataset using two different approaches:

- PGCNA : transposed matrices analysed with pgcna2 (-f 1, -e 3 -n 1000) with 1,000 clusterings, selecting the best clustering based on modularity score.
- Consensus clustering (CC): transposed matrices analysed using the R package ConsensusClusterPlus (maxK:14, linkage:average, algorithm:pam/HC, pItem:0.9, pFeatures:0.8, reps:100, distance:pearson)[7].

#### Assess cluster recurrence using machine learning

Different selections of genes/MEV/NEV, different clustering approaches (e.g. PGCNA, CC) and indeed just different clustering parameters will yield different results. Approaches like CC as part of their workflow will produce information about cluster stability giving an indication on how stably a cluster/module is resolved within that dataset. However, even if a set of clusters/modules is shown to be stable it does not mean that they can be recurrently found in other datasets. That is, it is possible that a set of stable clusters that exist in one dataset are never found in another dataset clustered using the same approach with the same parameters.

To address this question, we can use machine learning to try to gauge how recurrent clusters are between different datasets by:

- Unsupervised-Cluster each dataset independently using the same method/feature-sets (Clustering approaches).
- Compare each dataset with every other dataset:
    - Find shared features (genes, MEV/NEVs).
    - Train a ML tool on the unsupervised-clusters (those discovered by clustering).
    - Predict those clusters in every other dataset.
    - Compare ML predictions vs unsupervised-clusters:
        - Calculate significance of individual overlaps using Fisher's exact test
        - Calculate significance of maximal overlap:
            - Normalise all count matrices to the same size (total=250)
            - For each predicted class find maximum overlap with unsupervised-clusters.
            - Randomly shuffle (250x) same sets of predictions/unsupervised-clusters and use to calculate zScore/Pval of observed overlaps.
            - Scale result by normalized Shannon entropy of module sizes (0—1). This punishes results that only assign samples to a subset of the total available clusters).

A machine learning (ML) tool was created using the python Scikit-learn package [8], an MLPClassifier with the following hyperparameters (alpha:0.001, learning_rate_init:0.01, hidden_layer_sizes:200, max_iter:100, beta_1:0.9, beta_2:0.9) was trained on each gene/MEV/NEV set for each dataset and then used to classify every other dataset (excluding the Reddy and NCI-DLBCL dataset). An example of the overlap between unsupervised/supervised clustering is shown in Methods Fig 1.

For each gene/NEV/MEV set the significance of the overlaps was calculated between each of the 16 included DLBCL datasets. For PGCNA the single clustering per dataset were compared, for CC the datasets were compared at each K level (2—14). The resultant z-scores (calculated against random shuffled data) were then converted to a single value by taking the median across all results per gene/NEV/MEV set.

The results for each gene/NEV/MEV set clustered using CC or PGCNA are shown in Fig 4a, Table 3 (all results in ST5). This shows the following:

- Using the top 1000/5000 most variant genes for consensus clustering provides a good result. However, this is outperformed by the gene selection approaches based on the PGCNA network.
- The SelectG_MADS (gene selection without using edge info) results are worse than just using the top 1000/5000 most variant genes when used at the gene level. However, once genes are collapsed to NEV level then it outperforms the Top1000/5000 gene approaches.
- In all contexts collapsing genes to the NEV level outperforms analysis at the gene level. This is probably partially due to the improved ML training due to a simplified set of parameters. However, given that a goal is often to be able to recover the same clusters in new datasets this is a benefit.
- In all contexts the SelectG_NEV-MADM generates the best results when comparable parameters are used. Though the improvements over SelectG_NEV-CV are only marginal.

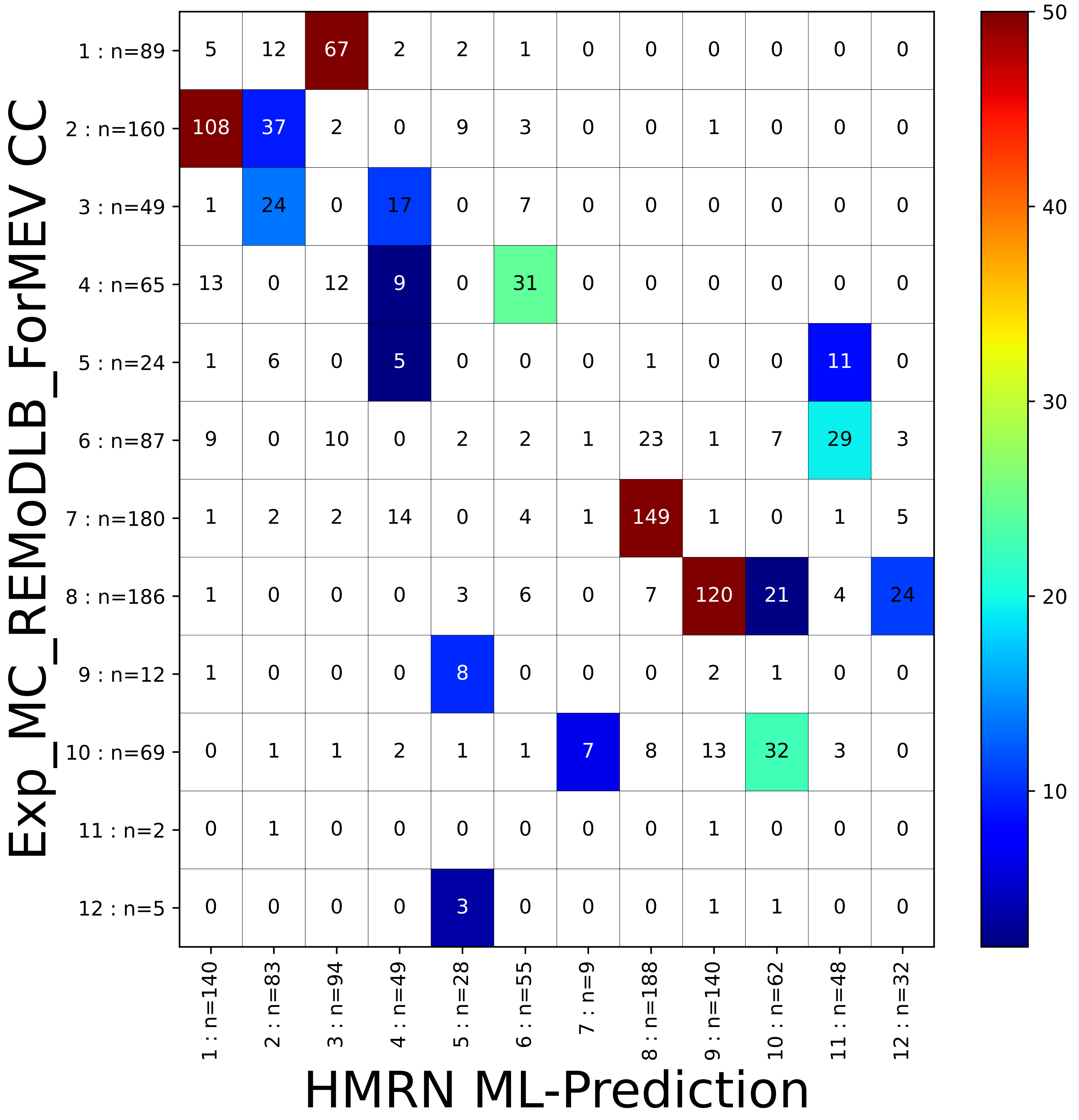


**Methods Fig 1. Recurrence of clusters within HMRN and REMoDLB dataset**
Shows the significance of the overlap between the ML classifier predictions for the REMoDLB dataset (trained on unsupervised HMRN clustering; x-axis) vs the unsupervised clustering of REMoDLB using the consensus cluster approach (y-axis). The overlaps are coloured by their -log10 p-values (Fisher's exact test; scale on the right). The number of overlapping samples are show in each square and the cluster number and number of samples belonging to that cluster is shown along each axis (e.g. 1 : n=140; shows cluster 1 that contains 140 samples, from the 928 samples in REMoDLB).

- In the context of DLBCL keeping most modules at the MEV level and using 2 key modules at the NEV level (Mod; Nbh M9M11) generates the best result. Outperforming splitting to the NEV level across all modules. This difference is larger in CC compared to PGCNA clusterings.
- In all CC results using the PAM method outperforms the default HC method. With the HC method larger K tend to result in a few very large clusters and the rest containing very few samples. The PAM methods splits the data into more equal subsets, and importantly these subsets are recurrently found across different datasets.

| Method | Gene Selection | Grouping Level | Collapse Level | Total Q1 | Total Q2 | Total Q3 | K10 | K11 | K12 | Comments | Chosen |
|---|---|---|---|---|---|---|---|---|---|---|---|
| Consensus Clustering (average_HC) | Top1000 | | Gene | 10.01 | 15.30 | 21.03 | 15.61 | 16.37 | 16.20 | Top 1000 most variant genes; excluding ChrY Gender Mod | |
| | Top5000 | | Gene | 9.16 | 13.89 | 18.48 | 13.95 | 14.08 | 14.17 | Top 5000 most variant genes; excluding ChrY Gender Mod | |
| | SelectG_MADS | Nbh | Gene | 10.12 | 15.07 | 20.53 | 14.94 | 15.30 | 15.70 | | |
| | SelectG_MADS | Nbh | NEV | 13.43 | 17.01 | 22.26 | 16.07 | 16.98 | 16.15 | | |
| | SelectG_NEV-CV | Nbh | Gene | 13.15 | 17.78 | 23.18 | 18.43 | 17.95 | 18.00 | | |
| | SelectG_NEV-CV | Nbh | NEV | 15.10 | 19.12 | 23.70 | 18.78 | 18.92 | 18.80 | | |
| | SelectG_NEV-MADM | Mod | Gene | 13.86 | 18.29 | 23.27 | 18.17 | 17.96 | 17.88 | Module Level | |
| | SelectG_NEV-MADM | Mod | MEV | 15.60 | 19.74 | 24.91 | 19.26 | 20.07 | 19.49 | Module Level | |
| | SelectG_NEV-MADM | Nbh | Gene | 11.17 | 15.25 | 19.79 | 15.29 | 16.48 | 15.59 | Less stringent cut-off (minDS0.2) | |
| | SelectG_NEV-MADM | Nbh | Gene | 12.92 | 17.54 | 22.52 | 16.76 | 18.81 | 17.76 | | |
| | SelectG_NEV-MADM | Nbh | NEV | 15.10 | 18.79 | 23.15 | 17.75 | 19.03 | 18.15 | | |
| | SelectG_NEV-MADM | Mod; Nbh (M9, M11) | Gene | 16.16 | 20.03 | 25.69 | 19.00 | 17.97 | 18.37 | Modules + Neighbourhoods M9M11 | |
| | SelectG_NEV-MADM | Mod; Nbh (M9, M11) | MEV/NEV | 19.25 | 22.68 | 28.45 | 21.32 | 20.58 | 20.29 | Modules + Neighbourhoods M9M11 | |
| Consensus Clustering (average_PAM) | Top1000 | | Gene | 12.93 | 17.38 | 21.64 | 18.07 | 17.57 | 17.78 | Top 1000 most variant genes; excluding ChrY Gender Mod | |
| | Top5000 | | Gene | 13.24 | 17.51 | 21.69 | 17.64 | 16.96 | 16.68 | Top 5000 most variant genes; excluding ChrY Gender Mod | |
| | SelectG_MADS | Nbh | Gene | 12.35 | 16.38 | 20.68 | 16.26 | 16.72 | 15.65 | | |
| | SelectG_MADS | Nbh | NEV | 16.37 | 19.89 | 24.03 | 19.34 | 18.85 | 18.17 | | |
| | SelectG_NEV-CV | Nbh | Gene | 15.15 | 18.77 | 22.76 | 18.42 | 18.44 | 18.58 | | |
| | SelectG_NEV-CV | Nbh | NEV | 19.18 | 22.55 | 26.53 | 22.15 | 21.75 | 22.34 | | |
| | SelectG_NEV-MADM | Mod | Gene | 16.00 | 19.79 | 24.13 | 20.02 | 19.74 | 19.75 | Module Level | |
| | SelectG_NEV-MADM | Mod | MEV | 19.34 | 23.23 | 27.52 | 22.82 | 22.78 | 22.27 | Module Level | |
| | SelectG_NEV-MADM | Nbh | Gene | 14.46 | 18.18 | 21.89 | 18.01 | 18.86 | 18.17 | Less stringent cut-off (minDS0.2) | |
| | SelectG_NEV-MADM | Nbh | Gene | 15.30 | 19.10 | 23.21 | 18.79 | 19.06 | 18.80 | | |
| | SelectG_NEV-MADM | Nbh | NEV | 19.50 | 22.74 | 27.09 | 22.82 | 21.99 | 22.43 | | |
| | SelectG_NEV-MADM | Mod; Nbh (M9, M11) | Gene | 16.52 | 20.54 | 24.95 | 19.00 | 18.28 | 18.07 | Modules + Neighbourhoods M9M11 | |
| | SelectG_NEV-MADM | Mod; Nbh (M9, M11) | MEV/NEV | 21.33 | 25.07 | 30.61 | 23.51 | 23.06 | 22.71 | Modules + Neighbourhoods M9M11 | + |
| PGCNA | SelectG_MADS | Nbh | Gene | 10.00 | 12.76 | 15.99 | | | | | |
| | SelectG_MADS | Nbh | NEV | 14.59 | 17.08 | 19.66 | | | | | |
| | SelectG_NEV-MADM | Mod | MEV | 15.08 | 17.80 | 20.75 | | | | Module Level | |
| | SelectG_NEV-CV | Nbh | NEV | 17.52 | 20.15 | 23.32 | | | | | |
| | SelectG_NEV-MADM | Nbh | NEV | 15.89 | 18.58 | 21.43 | | | | Less stringent cut-off (minDS0.2) | |
| | SelectG_NEV-MADM | Nbh | NEV | 18.04 | 21.23 | 24.29 | | | | | |
| | SelectG_NEV-MADM | Mod; Nbh (M9, M11) | MEV/NEV | 19.47 | 21.78 | 24.62 | | | | Modules + Neighbourhoods M9M11 | + |

Table 3: Machine learning assessment of cluster recurrence across different gene/MEV/NEV sets in both Consensus Clustering and PGCNA clustering of 16 DLBCL datasets. Table contains following columns Gene Selection : the gene/MEV/NEV set used (Selection of genes/MEV/NEV); Grouping Level: level at which genes are selected – Module (Mod), Neighbourhood (Nbh) or mixture (Mod; Nbh); Collapse Level : level at which genes are collapsed – gene (not collapsed), MEV (Module Expression Value), NEV (Neighbourhood Expression Value) or mixed (MEV/NEV); Total Q1/Q2/Q3: the median Z-score of significance of overlaps between all pairs of DLBCL datasets across all clusterings in quartiles 1—3; K10—K12: median Z-score of significance of overlaps at given CC K level; Comments : additional information; Chosen : the gene selection, grouping/collapse-level used for downstream analysis.

The results in Table 3 were used to select the best attributes (SelectG_NEV-MADM, Mod/Nbh M9M11) for downstream analysis.

### Lymphoma Communities (LC)

The HMRN CC/PGCNA communities from the chosen gene set (SelectG_NEV-MADM, Mod/Nbh M9M11) were each used to build a voting classifier with the python Scikit-learn package and the following hyperparameters:

- VotingClassifier (voting:soft):
  - KNeighborsClassifier (n_neighbors:5, weights:distance, algorithm:auto, leaf_size:100, metric:correlation)

- MLPClassifier (alpha:0.001, batch_size:50, learning_rate_init:0.001, hidden_layer_sizes:100, max_iter:100, beta_1:0.9, beta_2:0.9)
- SVC (kernel:rbf, degree:2, coef0:1, gamma:auto, C:5, class_weight:balanced)

The classifier was then used to predict the CC/PGCNA communities in the Reddy and REMoDLB datasets. A Mutation meta-analysis was carried out across the 3 datasets (HMRN, REMoDLB and Reddy), shown in Methods Fig 2. This shows that while both CC and PGCNA discover communities that have significant enrichment/depletion of mutations they capture some different features. For instance, PGCNA community 9 captures a BCL10/NOTCH2 association missing from the CC analysis and conversely CC communities 2/3 separate CARD11/EBF1 mutations from the SOCS1/SGK1 enriched GCB group. We thus combined the HMRN CC/PGCNA results by selecting significant overlaps ($p$-value < 0.0001) that form a community in either CC/PGCNA containing > 5 samples Methods Fig 3. This generated a high confidence set of Lymphoma Communities (LC; n=298) that formed a ML training dataset.

### Machine learning

To recover the LC in other datasets a machine learning (ML) tool was created using the python Scikit-learn package [8].

#### Input data

The CC/PGCNA overlapping communities in the HMRN data (see Lymphoma Communities (LC)) was used as a training set for machine learning. This consists of 298 samples x 41 MEV/NEV (Methods Fig 4a).

#### Training and validation datasets

The 298 HMRN dataset was split using the test_train_split function, stratifying on the LC class label, to give class balanced randomised training (n=238) and validation (n=60) datasets. The validation dataset was set aside to test the final selected model.

#### Model selection

The training data was used to carry out stratified 5-fold cross-validation across 7 different machine learning methods (see Methods Fig 5):

- ET: ExtraTrees
- GPC : GaussianProcessClassifier
- KNN : KNeighborsClassifier
- NN : MLPClassifier
- RF : RandomForestClassifier
- SVM : LinearSVC
- SVC : SVC

Using the GridSearchCV method 4,802 parameters were tested across the ML tools scoring with the Matthews correlation coefficient (MCC); Table 3.

| Type | Method | Parameters Tested | Max CrossVal MCC | Median MCC |
|---|---|---|---|---|
| Individual | ET | 27 | 0.80 | 0.78 |
| | GPC | 240 | 0.83 | 0.79 |
| | KNN | 2,240 | 0.86 | 0.62 |
| | NN | 1,728 | 0.87 | 0.83 |
| | RF | 270 | 0.81 | 0.74 |
| | SVM | 72 | 0.76 | 0.67 |
| | SVC | 225 | 0.88 | 0.81 |
| | Total | 4,802 | | |
| Voting | KNN, NN, SVC | Soft Vote | 0.89 | |

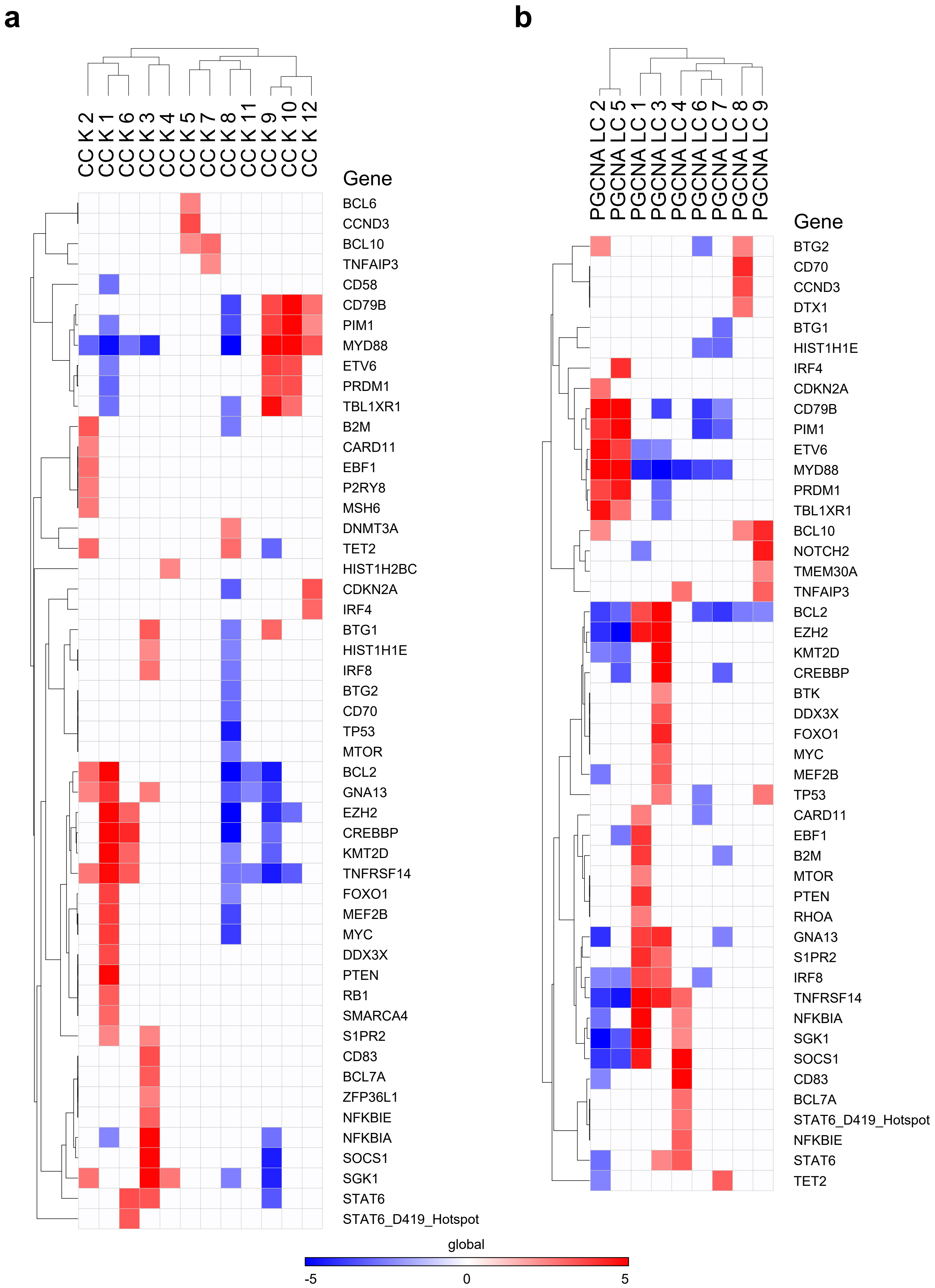


**Methods Fig 2. Consensus Clustering and PGCNA association with mutations**

Association of CC/PGCNA with mutation status. The differential enrichment of gene mutations for (**a**) CC K12 communities and (**b**) PGCNA communities across three datasets. Shown is combined significance (Stouffer method) of community enrichment/depletion of mutations as z-scores on a blue (significant depletion) to red (significant enrichment) scale (-5 to +5). X-axis shows hierarchically clustered community and y-axis gene symbols. Only mutations with p-value < 0.01, occurring in ≥ 2 datasets were retained, Z-scores for p-values > 0.01 were set to 0.

Table 3: Method – machine learning tool tested, Parameters Tested – the number of parameters tested in the grid search, Max CrossVal MCC – maximum mean MCC across the 5-fold stratified cross-validation, Median MCC – median of the mean MCC across the parameters tested.

The best 3 models, using their optimal parameters, were combined using a soft voting approach to give a model with a mean MCC of 0.89 across the 5-folds. This model was then tested on the unseen validation data (n=60) where it achieved an MCC of 0.87 (Methods Fig 6). This final model, termed ML_LC, was retrained using all 298 training samples; this model was used for all classifications.

#### Classification of datasets

The ML_LC classifier was first used to reclassify the total HMRN dataset (n=451; including the samples removed from the select training data; Methods Fig 4b), thus assigning a p-value for each LC to each sample. 6 DLBCL datasets (GSE10846, GSE31312, HMRN, NCI-DLBCL, Reddy, REMoDLB) were each converted to median MEV/NEV values and then classified using the ML_LC classifier into the 12 LC (See EDF5, EDF6).

### Statistical analyses

#### Gene signature data and enrichment analysis

A dataset of 20,707 gene signatures was created by merging signatures downloaded from http://lymphochip.nih.gov/signaturedb/ (SignatureDB), http://www.broadinstitute.org/gsea/msigdb/index.jsp MSigDB V7.4 (MSigDB C1--C8 and H; excluding C5), Human CORUM complexes with > 2 genes (http://mips.helmholtz-muenchen.de/corum/#download), UniProt keywords (parsed XML; http://www.uniprot.org/downloads) and 18 papers (PMIDs:12975453,15550490,19412164,20725040,21179087,23563690,23584089,23584090,23700391,23871637,24138885,24220563,24336226,24644022,25800755,25822800,28735890, 32603407). A gene ontology gene-set was created using an in-house python script. This parses a gene association file (http://geneontology.org/page/download-annotations ) to link genes with ontology terms, then uses the ontology structure (.obo file; http://geneontology.org/docs/download-ontology/) to propagate these terms up to the root. The resultant gene-set contained 22,865 terms.

The gene-ontology and gene-signatures sets were merged to give a final signature set of 43,572 terms. Enrichment of modules for signatures was assessed using a hypergeometric test, where the draw is the module genes, the successes are the signature genes, and the population is the genes present on the platform.

### Survival analysis

#### Gene/LC level meta-analysis

The Survival library for R was used to analyze right-censored survival data for the data sets where this was available. Within each data set the expression of each gene (as z-score) or the ML_LC LC p-value was used as a continuous variable in a Cox Proportional Hazards model and the ML_LC community for a Kaplain-Meier estimator (using merged survival data). Across data sets a meta-analysis was conducted by fitting a fixed-effect model (R metafor package) to the hazard ratios, weighted by data set size. rma(yi=lnHazardRatio,sei=standardErr,weights=dataSetSize,weighted=TRUE,method="FE")[9]. Kaplain-Meier survival plots were generated using the R survminer ggsurvplot function, showing pvalues from the log-rank test.

### Mutation analysis

Analysis of the correlation between mutations and other features was carried out for three datasets using HMRN driver mutations (188 genes for 431 samples), REMoDLB mutations (70 genes for 400 samples)

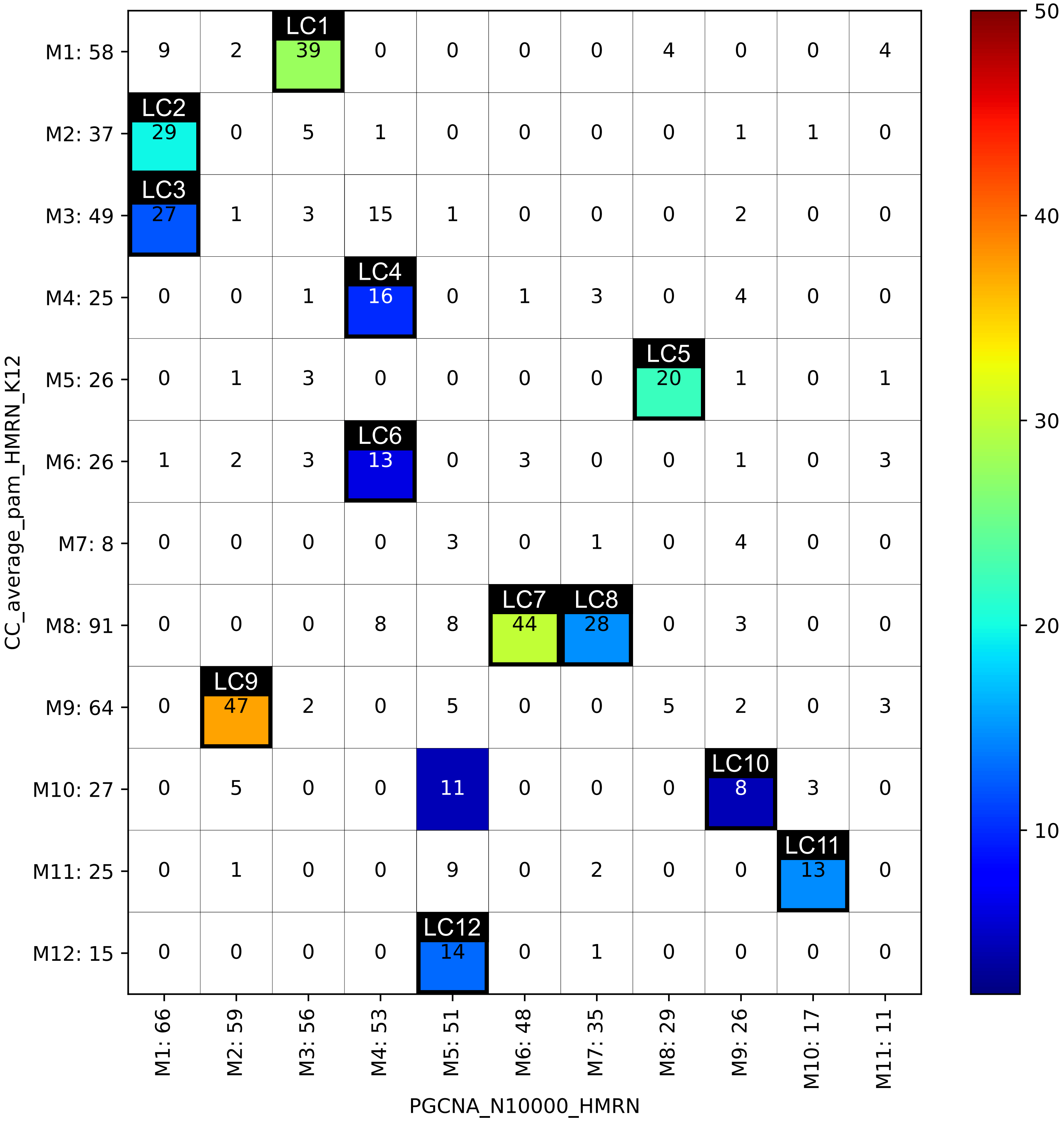


**Methods Fig 3. Lymphoma communities (LC) from overlap of Consensus Clustering and PGCNA**
The generation of the Lymphoma Communities based on the overlaps between Consensus Clustering (CC) and PGCNA in the HMRN dataset. Shows the significance of the overlap between the CC (y-axis) and PGCNA (x-axis) communities. The overlaps are coloured by their -log10 *p*-values (Fisher's exact test; scale on the right). The number of overlapping samples are show in each square and the community number and number of samples belonging to that cluster is shown along each axis. The overlapping samples representing each Lymphoma Community (LC) are highlighted with bold boxes. The LC samples formed the HMRN training data (n=298) used for machine learning development. Overlaps with *p*-values > 0.0001 were set to white.

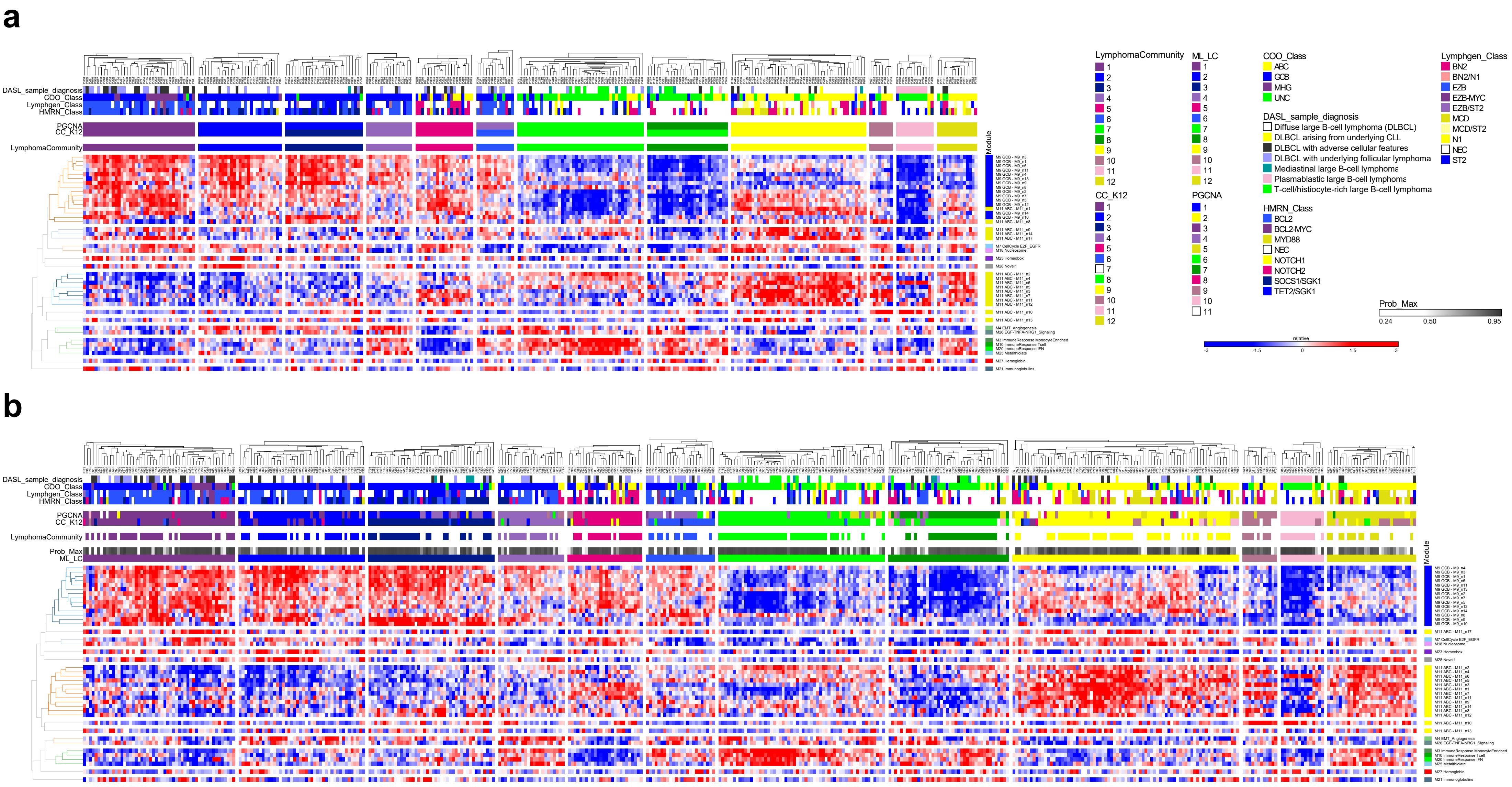


**Methods Fig 4. HMRN training and classified data**

HMRN DLBCL dataset was used to derive consistent communities of DLBCL cases based on CC/PGCNA clustering using 41 MEV/NEVs. Twelve Lymphoma Communities (LC) were resolved based on communities shared between CC/PGCNA (Methods Fig.3) and used to train a machine learning tool (ML_LC). (**a**) the HMRN training data cases hierarchically clustered within each LC group (LymphomaCommunity; n=298). (**b**) HMRN data hierarchically clustered withing each assigned LC (ML_LC; n=451). MEV and NEV are shown on a blue (low) to red (high) z-score colour scale. Modules and neighbourhoods are separated across the y-axis and colour coded for GCB (blue) and ABC (yellow) neighbourhoods. Meta-data is provided (top right) showing – **ML_LC**: the LC assigned by the ML_LC classifier, **Prob_Max**: the ML_LC classification confidence (white == 0.5, to dark-grey == 0.95), **LymphomaCommunity**: the training data LC groups, **CC_K12**: the CC communities, **PGCNA**: the PGCNA communities, **HMRN_Class**: the HMRN mutation classifications (PMID:32187361), **Lymphgen_Class**: assigned LymphGen classifications (PMID:32289277), **COO_Class**: the cell-of-origin/MHG class, **DASL_sample_diagnosis**: the morphological classification.

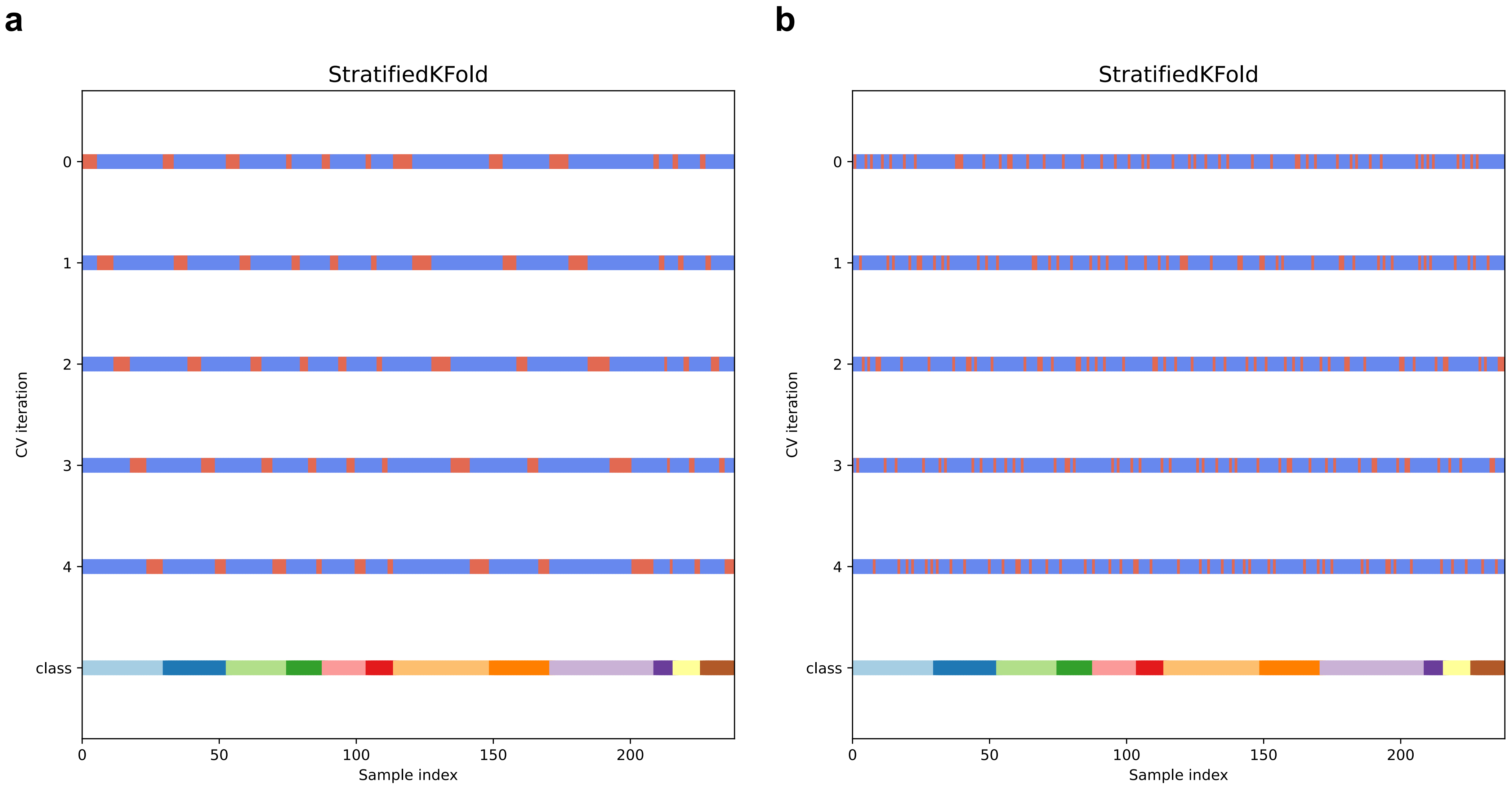


**Methods Fig 5. K-Fold Statification**

(**a**) Shows stratified K-folds, preserving the percentage of samples from each class. For each of the 5 folds the training data is shown in blue and the test in red. The Integrated Context Classifications are shown as the class underneath the iterations.

(**b**) Shows the same as (a), with a stratification of the samples chosen at each K-fold. This fixed stratification of the data was used for all model testing.

**a**

**Training — Perfect**

| | LC 1 | 2 | 3 | 4 | 5 | 6 | 7 | 8 | 9 | 10 | 11 | 12 |
|---|---|---|---|---|---|---|---|---|---|---|---|---|
| 1 | 31 | 0 | 0 | 0 | 0 | 0 | 0 | 0 | 0 | 0 | 0 | 0 |
| 2 | 0 | 23 | 0 | 0 | 0 | 0 | 0 | 0 | 0 | 0 | 0 | 0 |
| 3 | 0 | 0 | 22 | 0 | 0 | 0 | 0 | 0 | 0 | 0 | 0 | 0 |
| 4 | 0 | 0 | 0 | 13 | 0 | 0 | 0 | 0 | 0 | 0 | 0 | 0 |
| 5 | 0 | 0 | 0 | 0 | 16 | 0 | 0 | 0 | 0 | 0 | 0 | 0 |
| 6 | 0 | 0 | 0 | 0 | 0 | 10 | 0 | 0 | 0 | 0 | 0 | 0 |
| 7 | 0 | 0 | 0 | 0 | 0 | 0 | 35 | 0 | 0 | 0 | 0 | 0 |
| 8 | 0 | 0 | 0 | 0 | 0 | 0 | 0 | 22 | 0 | 0 | 0 | 0 |
| 9 | 0 | 0 | 0 | 0 | 0 | 0 | 0 | 0 | 38 | 0 | 0 | 0 |
| 10 | 0 | 0 | 0 | 0 | 0 | 0 | 0 | 0 | 0 | 7 | 0 | 0 |
| 11 | 0 | 0 | 0 | 0 | 0 | 0 | 0 | 0 | 0 | 0 | 10 | 0 |
| 12 | 0 | 0 | 0 | 0 | 0 | 0 | 0 | 0 | 0 | 0 | 0 | 11 |

**Training — ML Model**

| | LC 1 | 2 | 3 | 4 | 5 | 6 | 7 | 8 | 9 | 10 | 11 | 12 |
|---|---|---|---|---|---|---|---|---|---|---|---|---|
| 1 | 30 | 0 | 1 | 0 | 0 | 0 | 0 | 0 | 0 | 0 | 0 | 0 |
| 2 | 2 | 20 | 1 | 0 | 0 | 0 | 0 | 0 | 0 | 0 | 0 | 0 |
| 3 | 0 | 1 | 20 | 0 | 1 | 0 | 0 | 0 | 0 | 0 | 0 | 0 |
| 4 | 0 | 0 | 0 | 13 | 0 | 0 | 0 | 0 | 0 | 0 | 0 | 0 |
| 5 | 0 | 0 | 0 | 0 | 15 | 0 | 0 | 0 | 1 | 0 | 0 | 0 |
| 6 | 1 | 1 | 0 | 0 | 0 | 5 | 3 | 0 | 0 | 0 | 0 | 0 |
| 7 | 0 | 0 | 0 | 0 | 0 | 0 | 31 | 4 | 0 | 0 | 0 | 0 |
| 8 | 0 | 0 | 0 | 0 | 0 | 0 | 3 | 18 | 0 | 0 | 1 | 0 |
| 9 | 1 | 0 | 0 | 0 | 0 | 0 | 0 | 0 | 37 | 0 | 0 | 0 |
| 10 | 0 | 0 | 0 | 0 | 0 | 0 | 0 | 0 | 0 | 7 | 0 | 0 |
| 11 | 0 | 0 | 0 | 0 | 0 | 0 | 0 | 0 | 1 | 0 | 9 | 0 |
| 12 | 0 | 0 | 0 | 0 | 0 | 0 | 0 | 1 | 1 | 0 | 0 | 9 |

MCC : 0.89

**b**

**Validation — Perfect**

| | LC 1 | 2 | 3 | 4 | 5 | 6 | 7 | 8 | 9 | 10 | 11 | 12 |
|---|---|---|---|---|---|---|---|---|---|---|---|---|
| 1 | 8 | 0 | 0 | 0 | 0 | 0 | 0 | 0 | 0 | 0 | 0 | 0 |
| 2 | 0 | 6 | 0 | 0 | 0 | 0 | 0 | 0 | 0 | 0 | 0 | 0 |
| 3 | 0 | 0 | 5 | 0 | 0 | 0 | 0 | 0 | 0 | 0 | 0 | 0 |
| 4 | 0 | 0 | 0 | 3 | 0 | 0 | 0 | 0 | 0 | 0 | 0 | 0 |
| 5 | 0 | 0 | 0 | 0 | 4 | 0 | 0 | 0 | 0 | 0 | 0 | 0 |
| 6 | 0 | 0 | 0 | 0 | 0 | 3 | 0 | 0 | 0 | 0 | 0 | 0 |
| 7 | 0 | 0 | 0 | 0 | 0 | 0 | 9 | 0 | 0 | 0 | 0 | 0 |
| 8 | 0 | 0 | 0 | 0 | 0 | 0 | 0 | 6 | 0 | 0 | 0 | 0 |
| 9 | 0 | 0 | 0 | 0 | 0 | 0 | 0 | 0 | 9 | 0 | 0 | 0 |
| 10 | 0 | 0 | 0 | 0 | 0 | 0 | 0 | 0 | 0 | 1 | 0 | 0 |
| 11 | 0 | 0 | 0 | 0 | 0 | 0 | 0 | 0 | 0 | 0 | 3 | 0 |
| 12 | 0 | 0 | 0 | 0 | 0 | 0 | 0 | 0 | 0 | 0 | 0 | 3 |

**Validation — ML Model**

| | LC 1 | 2 | 3 | 4 | 5 | 6 | 7 | 8 | 9 | 10 | 11 | 12 |
|---|---|---|---|---|---|---|---|---|---|---|---|---|
| 1 | 8 | 0 | 0 | 0 | 0 | 0 | 0 | 0 | 0 | 0 | 0 | 0 |
| 2 | 0 | 6 | 0 | 0 | 0 | 0 | 0 | 0 | 0 | 0 | 0 | 0 |
| 3 | 0 | 0 | 5 | 0 | 0 | 0 | 0 | 0 | 0 | 0 | 0 | 0 |
| 4 | 0 | 0 | 1 | 1 | 0 | 1 | 0 | 0 | 0 | 0 | 0 | 0 |
| 5 | 1 | 0 | 0 | 0 | 2 | 0 | 0 | 0 | 1 | 0 | 0 | 0 |
| 6 | 0 | 1 | 0 | 0 | 0 | 2 | 0 | 0 | 0 | 0 | 0 | 0 |
| 7 | 0 | 0 | 0 | 0 | 0 | 0 | 8 | 1 | 0 | 0 | 0 | 0 |
| 8 | 0 | 0 | 0 | 0 | 0 | 0 | 0 | 6 | 0 | 0 | 0 | 0 |
| 9 | 0 | 0 | 0 | 0 | 0 | 0 | 0 | 0 | 8 | 0 | 0 | 1 |
| 10 | 0 | 0 | 0 | 0 | 0 | 0 | 0 | 0 | 0 | 1 | 0 | 0 |
| 11 | 0 | 0 | 0 | 0 | 0 | 0 | 0 | 0 | 0 | 0 | 3 | 0 |
| 12 | 0 | 0 | 0 | 0 | 0 | 0 | 0 | 0 | 0 | 0 | 0 | 3 |

MCC : 0.87

**Methods Fig 6. Confusion-matrix for the ML_LC classifier**

(**a**) The confusion matrices for the ML_LC model trained on the entire training dataset (n=267). The top confusion matrix (Perfect) shows the confusion matrix if all cases are classified correctly, the bottom confusion matrix (ML Model) shows the incorrect classifications as numbers horizontally shifted from the diagonal, the column showing which class they have being incorrectly assigned to. The Matthews Correlation Coefficient (MCC) is shown at the bottom.

(**b**) Same as for (**a**), with the ML_LC model tested on the held-out validation data (n=70) and Reddy mutations (150 genes for 624 samples)[10–12]. The mutations were converted to a binary matrix for downstream analysis.

#### MEV/NEV analysis

For each dataset the point-biserial correlations were calculated between all pairs of mutated gene and MEV/NEV. The resulting correlation p-values were converted to z-scores (python stats.norm.ppf) to convey the ± correlation along with its significance.

#### LC analysis

The significance of the overlap between mutations and LC was calculated using a hypergeometric test, where the draw is the samples in an LC, the successes are the number of mutations of a gene in the LC and the population is total number of mutations of that gene.

#### Mutation meta-analysis

The individual MEV/LC mutation analyses were combined to give a single meta result each. In both cases the p-values were combined using the python scipy.stats combine_pvalues function (stouffer method). The correlation (MEVs) or enrichment/depletion (LC) states were used to check agreement of the direction of effect across the datasets. If there was a disagreement then the mode was used, merging only the p-values for the datasets in agreement. The Z-scores of the combined pvalues (from scipy.stats combine_pvalues; stouffer Z) were used to convey the ± correlation/enrichment along with its significance. A matrix was output for all mutations that had a combined p-value < 0.01, occurring in ≥ 2 datasets (LC: Fig 5a, MEV: EDF4).

### LC differential gene expression

The genes differentially expressed between each pair of LC was calculated for each dataset using the R Limma package[13]. The results were combined across datasets using the python scipy.stats combine_pvalues function (stouffer method). The results are available at https://mcare.link/DLBCL.

### Data visualisations

#### LC visualisations

The expression patterns of the LC were visualised at the MEV/NEV level in the HMRN dataset (Fig 4b) and for the other 5 classified datasets (EDF5). A simplified view was generated by calculating the Inter Quartile Range (IQR) for each module/neighbourhood MEV for each LC (EDF6), the median from which was visualised at the network level for the genes present on the HMRN platform (EDF7 and EDF8).

#### Heatmaps

All heatmap visualisations were generated using the Broad GENE-E package (https://software.broadinstitute.org/GENE-E/) hierarchically clustering (Pearson correlations and average linkage) and displayed along with available meta data.

#### Violin plots

Violin plots were generated using the R package ggplot2 and show the median (blue square) along with IQR (Fig1d). Violin plots in Fig 4a were generated with the Python seaborn package, displaying the Q1—Q3 along with a swarmplot displaying the individual results.

### Software and data

The DLBCL LC classifier is available along with networks and analysis results at https://mcare.link/DLBCL.

#### Data processing

All analyses were undertaken on ARC4, part of the High-Performance Computing facilities at the University of Leeds, UK.
